# Genetic predisposition to altered blood cell homeostasis is associated with glioma risk and survival

**DOI:** 10.1101/2023.10.15.23296448

**Authors:** Linda Kachuri, Geno A. Guerra, George A. Wendt, Helen M. Hansen, Annette M. Molinaro, Paige Bracci, Lucie McCoy, Terri Rice, John K. Wiencke, Jeanette E. Eckel-Passow, Robert B. Jenkins, Margaret Wrensch, Stephen S. Francis

**Affiliations:** Department of Epidemiology & Population Health, Stanford University School of Medicine, Stanford, CA; Stanford Cancer Institute, Stanford University School of Medicine, Stanford, CA; Department of Neurological Surgery, University of California San Francisco, San Francisco, CA; Department of Epidemiology & Biostatistics, University of California San Francisco, San Francisco, CA; Weill Institute for Neurosciences, University of California San Francisco, San Francisco, US; Division of Biomedical Statistics and Informatics, Mayo Clinic, Rochester, Minnesota, USA; Department of Laboratory Medicine and Pathology, Mayo Clinic, Rochester, Minnesota, USA

## Abstract

Glioma is a highly fatal brain tumor comprised of molecular subtypes with distinct clinical trajectories. Observational studies have suggested that variability in immune response may play a role in glioma etiology. However, their findings have been inconsistent and susceptible to reverse causation due to treatment effects and the immunosuppressive nature of glioma. We applied genetic variants associated (p<5×10^−8^) with blood cell traits to a meta-analysis of 3418 glioma cases and 8156 controls. Genetically predicted increase in the platelet to lymphocyte ratio (PLR) was associated with an increased risk of glioma (odds ratio (OR)=1.25, p=0.005), especially in IDH-mutant (IDH_mut_ OR=1.38, p=0.007) and IDH_mut_ 1p/19q non-codeleted (IDH_mut-noncodel_ OR=1.53, p=0.004) tumors. However, reduced glioma risk was observed for higher counts of lymphocytes (IDH_mut-noncodel_ OR=0.70, p=0.004) and neutrophils (IDH_mut_ OR=0.69, p=0.019; IDH_mut-noncodel_ OR=0.60, p=0.009), which may reflect genetic predisposition to enhanced immune-surveillance. In contrast to susceptibility, there was no association with survival in IDH_mut-noncodel;_ however, in IDH_mut_ 1p/19q co-deleted tumors, we observed higher mortality with increasing genetically predicted counts of lymphocytes (hazard ratio (HR)=1.65, 95% CI: 1.24-2.20), neutrophils (HR=1.49, 1.13-1.97), and eosinophils (HR=1.59, 1.18-2.14). Polygenic scores for blood cell traits were also associated with tumor immune microenvironment features, with heterogeneity by IDH status observed for 17 signatures related to interferon signaling, PD-1 expression, and T-cell/Cytotoxic responses. In summary, we identified novel, immune-mediated susceptibility mechanisms for glioma with potential disease management implications.

**SIGNIFICANCE:** This study suggests that genetic determinants of peripheral blood cell counts influence subtype-specific glioma susceptibility and mortality, with potential effects on the tumor immune microenvironment. These findings may have disease management implications.

## INTRODUCTION

Glioma is the most common malignant central nervous system tumor in the United States and globally^1,2^. Despite being a relatively rare primary cancer, gliomas make an outsize contribution to cancer mortality due to limited advances in treatment. Identification of molecular subtypes, which have been incorporated into the World Health Organization (WHO) brain tumor classification scheme since 2016, has helped delineate gliomas with distinct clinical presentation and survival trajectories beyond traditional histopathologic categories. Two major molecular features used to classify gliomas are mutations in *IDH1* (isocitrate dehydrogenase 1) or *IDH2* genes, collectively referred to as IDH mutations, and chromosome 1p and 19q co-deletion status^3,4^. Patients with IDH-mutant gliomas have a more favorable prognosis, but these tumors tend to progress, recur as higher grades, and become resistant to therapy. IDH wildtype tumors are the most aggressive, with a median survival of only 1.2 years, compared to 17.5 years for patients with IDH mutation and 1p/19q co-deletion^4,5^.

There are few non-genetic risk factors for glioma and even less is known about differences in putative risk factors across molecular subtypes. Exposure to ionizing radiation is the only established causal factor, although it accounts for a low proportion of glioma cases at the population level^4^. An accumulation of findings from observational studies makes a compelling case for an immune component in glioma etiology. History of infection with varicella zoster virus (VZV), a neurotropic α-herpesvirus that causes chickenpox and shingles^6,7^, and the presence of IgG antibodies to VZV^7^ have been inversely associated with glioma risk. This has recently extended to prognosis, where stronger VZV reactivity was associated with improved survival^8^. History of allergies or other atopic conditions (hay fever, eczema, and asthma)^9-12^ and increased IgE levels^13,14^ have been associated with a decreased risk of glioma in multiple observational studies. However, an analysis using genetic variants associated with atopic conditions and IgE levels did not support these findings and detected only inconsistent protective effects for atopic dermatitis^15^. Other genetic association studies have uncovered protective effects of stronger immune reactivity to viral infection^16^ and an inverse relationship between genetic susceptibility to glioma and autoimmune conditions^17^.

The mechanisms underlying these associations with immune function remain elusive. One candidate hypothesis for the observed protective effects relates to heightened immune activation and improved immunosurveillance that translates into enhanced tumor clearance^4,10,13^. Commonly measured hematologic indices, such as counts of white blood cells and platelets, reflect systemic immune and inflammatory responses and are frequently dysregulated in cancer patients. Immune dysregulation is well documented in glioma patients^18-20^. Low proportions of CD4 T-cells have been linked to diminished survival^5,21^. While these observations may be relevant for prognostic stratification, the potential etiologic role of alterations in immune cells profiles is confounded by effects of dexamethasone treatment and systemic changes caused by the disease process^20^.

In this study, we leverage a large collection of glioma cases with genome-wide genetic data and information on molecular subtypes to examine how genetic predisposition to increased counts of leukocytes and platelets relate to glioma risk and survival. Genome-wide association studies (GWAS) have demonstrated overlap between genetic loci involved in regulation of blood cell traits and cancer susceptibility^22-24^. Furthermore, host immune responses to infection and cancer share important, evolutionarily conserved, pathways, many of which are dependent on platelet- and leukocyte-driven processes. By relying on germline determinants of constitutive variation in blood cell counts, Mendelian randomization (MR) provides associations that are not affected by reverse causation, a major limitation of observational studies due to alterations in immune cell frequencies caused by tumorigenesis and treatment. In addition to investigating glioma susceptibility, we also comprehensively characterize the role of genetically predicted immune cell frequencies with overall glioma survival.

## METHODS

### Study population

We assembled data from three case-control studies with available molecular and genetic data to facilitate analyses stratified by glioma subtypes, as described in Guerra et al^16^. Briefly, we analyzed 1973 cases from the Mayo Clinic and University of California San Francisco (UCSF) Adult Glioma Study (AGS) with 1859 controls from the Glioma International Case-Control Study (GICC), an additional 659 cases and 586 controls from the UCSF AGS, and 786 glioma cases from The Cancer Genome Atlas (TCGA) with 5711 controls from the Wellcome Trust Case Control Consortium (WTCCC)^3,25-27^. All participants were over 18 years of age. Imputation was performed using the TOPMed reference panel. Analyses were restricted to individuals of predominantly (>70%) European genetic ancestry. Study-specific GWAS results were meta-analyzed for a total sample size of 3418 cases and 8156 controls **(Figure S1)**. As our analyses are based on historic cohort data, we lack the full set of molecular markers required to align our case definitions with WHO 2021 glioma classifications. However, using the available information we delineated prognostically significant phenotypes that distinguished IDH mutated (IDH_mut_), IDH mutated 1p/19q co-deleted (IDH_mut-codel_), IDH mutated without 1p/19q co-deletion (IDH_mut-nondel_), and IDH wildtype (IDH_wt_) tumors. Collection of patient samples and associated clinicopathological information was undertaken with written informed consent and ethical board approval was obtained from the UCSF Committee on Human Research (USA) and the Mayo Clinic Office for Human Research Protection (USA).

### Mendelian randomization and colocalization

Mendelian randomization (MR) is a statistical method that can infer causal exposure-outcome relationships under a strict set of assumptions by using genetic variants strongly associated with the exposure. The basic MR framework involves fitting a weighted linear regression between effect sizes for the outcome and effect sizes for the exposure. If all instruments are valid, the resulting slope provides an estimate of the causal effect of the exposure. Genetic instruments for blood cell traits were developed in the UK Biobank (UKB) cohort, as described in Kachuri et al.^22^, and applied to the glioma GWAS meta-analysis (3418 cases and 8156 controls). Briefly, two-stage GWAS in cancer-free controls of predominantly European ancestry was followed by Multi-Trait Analysis of GWAS (MTAG)^28^. Genetic instruments for blood cell traits were selected from MTAG results based on the following criteria: linkage disequilibrium (LD) *r*^2^<0.05 with a 10 Mb window with discovery *P*<5×10^−8^ and replication *P*<0.05.

A fundamental, but untestable, MR assumption of “no horizontal pleiotropy” requires that instruments influence the outcome exclusively through the exposure of interest, without any direct effects on risk. Since this assumption cannot be verified, the sensitivity of the observed results to different scenarios of pleiotropy were assessed by examining consistency across multiple MR estimators. Maximum likelihood (ML) is the most restrictive method, unbiased only when all instruments are valid. Inverse variance weighted multiplicative random-effects (IVW-mre) approach accounts for heterogeneity caused by non-directional pleiotropy^29,30^. MR Egger was reported if directional horizontal pleiotropy was indicated by the deviation of the regression intercept from 0 (p<0.05). Weighted median (WM)^31^ allows up to 50% of the weights to be contributed by invalid instruments. MR RAPS (Robust Adjusted Profile Score)^32,33^ employs a robust loss function to limit the influence of outlier instruments. MR pleiotropy residual sum and outlier (PRESSO)^34^ identifies and corrects for instruments driving the difference between the observed residual sum of squares and the expectation, simulated under the null hypothesis of no horizontal pleiotropy. For each MR estimator, odds ratios (OR) and 95% confidence intervals (CI) correspond to a genetically predicted standard deviation (SD) increase in lymphocytes, monocytes, neutrophils, basophils, and eosinophils. For blood cell ratios, effects were estimated per one-unit increase.

Given the correlated nature of blood cell traits, multivariable MR was performed to estimate conditional effects on glioma risk by simultaneously regressing all instruments across all exposures. As in univariate MR, weighting was based on the inverse of the variance of outcome effect sizes (MV-IVW). We also performed regularization-based MVMR, where the penalty controlling level of sparsity is tuned based on heterogeneity (MV-LASSO)^35^.

We also fine-mapped associations at genetic instruments for blood cell traits that also reached genome-wide significance (*P*<5×10^−8^) in for at least one glioma subtype in our data. Colocalization was performed in a 100-bp region centered on the risk variant to estimate the posterior probability (PP) of a shared causal signal for disease susceptibility and regulation of blood cell homeostasis. Colocalization analyses were performed using HyPrColoc for genetic variants that were associated with multiple candidate mechanisms.

### Genetic associations with survival

We explored associations between genetically predicted blood cell profiles and survival in glioma patients by fitting a polygenic score (PGS) for each trait computed as a weighted sum of trait-increasing alleles:

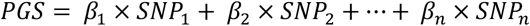

with weights (β) corresponding to the same effect sizes as those used in the MR analysis. Hazard ratios (HR) per SD increase in the standardized PGS were estimated using a Cox proportional hazards model with adjustment for age, sex, the top 10 principal components (PC) of genetic ancestry, and study site for the combined AGS/Mayo Clinic case series. Follow-up time was calculated from the date of first surgery to either date of death or date of last known contact. Study-specific PGS associations were meta-analyzed.

Significant PGS associations were followed up in Mendelian randomization analyses, however, assessing causality for disease progression endpoints may be complicated by index event bias^36,37^. Selection based on disease status in case-only analyses may introduce bias into genetic associations with progression because cancer risk factors may become correlated when restricting to cases, which in turn may induce associations between risk factors and cancer progression events^37^. We applied the Dudbridge et al.^37^ method to 41,528 pruned variants (LD *r*^2^<0.10 in 250 kb windows) with MAF ≥0.05 and INFO>0.90 to test for the presence of index event bias and estimate the corresponding correction factor (+). This approach regresses SNP effects on risk against their effects on survival^37^. Bias-corrected effects on survival are estimated as follows: 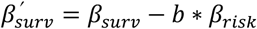 where β_*risk*_ is obtained from the case-control glioma GWAS.

### Genetic associations with the tumor immune microenvironment features

We explored several potential mechanisms through which genetic determinants of blood cell traits may be influencing glioma susceptibility and progression. First, we examined associations between selected blood trait PGS and heritable tumor immune microenvironment features identified by Sayaman et al^38^ in TCGA. Of the 33 pan-cancer immune phenotypes, 28 were available for analysis in glioma subjects.

## RESULTS

### Genetically inferred blood cell profiles and glioma risk

Summary statistics were available from a GWAS meta-analysis of 3418 glioma cases with available molecular profiling and 8156 controls. The majority of cases (n=1479) had IDH wildtype (IDH_wt_) tumors. A total of 1074 cases harbored somatic *IDH1*/*IDH2* mutations (IDH_mut_), of which 396 were also 1p/19q co-deleted (IDH_mut-codel_) while 622 had 1p/19q non-codeleted (IDH_mut-noncodel_).

Analysis of glioma overall identified a susceptibility signal for platelet to lymphocyte ratio (PLR) **(Figure 1; Supplementary Table S1)**, suggesting that genetic predisposition to an increase in the relative abundance of platelets to lymphocytes confers an increased glioma risk (OR_ML_=1.25, 95% CI: 1.10-1.43, p=9.1×10^−4^). These results persisted in sensitivity analyses (OR_IVW-MRE_=1.25, p=4.9×10^−3^; OR_PRESSO_=1.21, p=8.8×10^−3^; OR_RAPS_=1.22, p=0.011). No consistent associations were observed for individual cell counts, although there was evidence of directional pleiotropy (Egger intercept β_0_≠0, p<0.05) for eosinophils, lymphocytes, monocytes, and platelets **(Supplementary Tables S2)**. Accounting for this with MR Egger detected an inverse association with glioma for eosinophil counts (OR_Egger_ = 0.67, 0.46-0.99, p=0.044).

**Figure 1:**
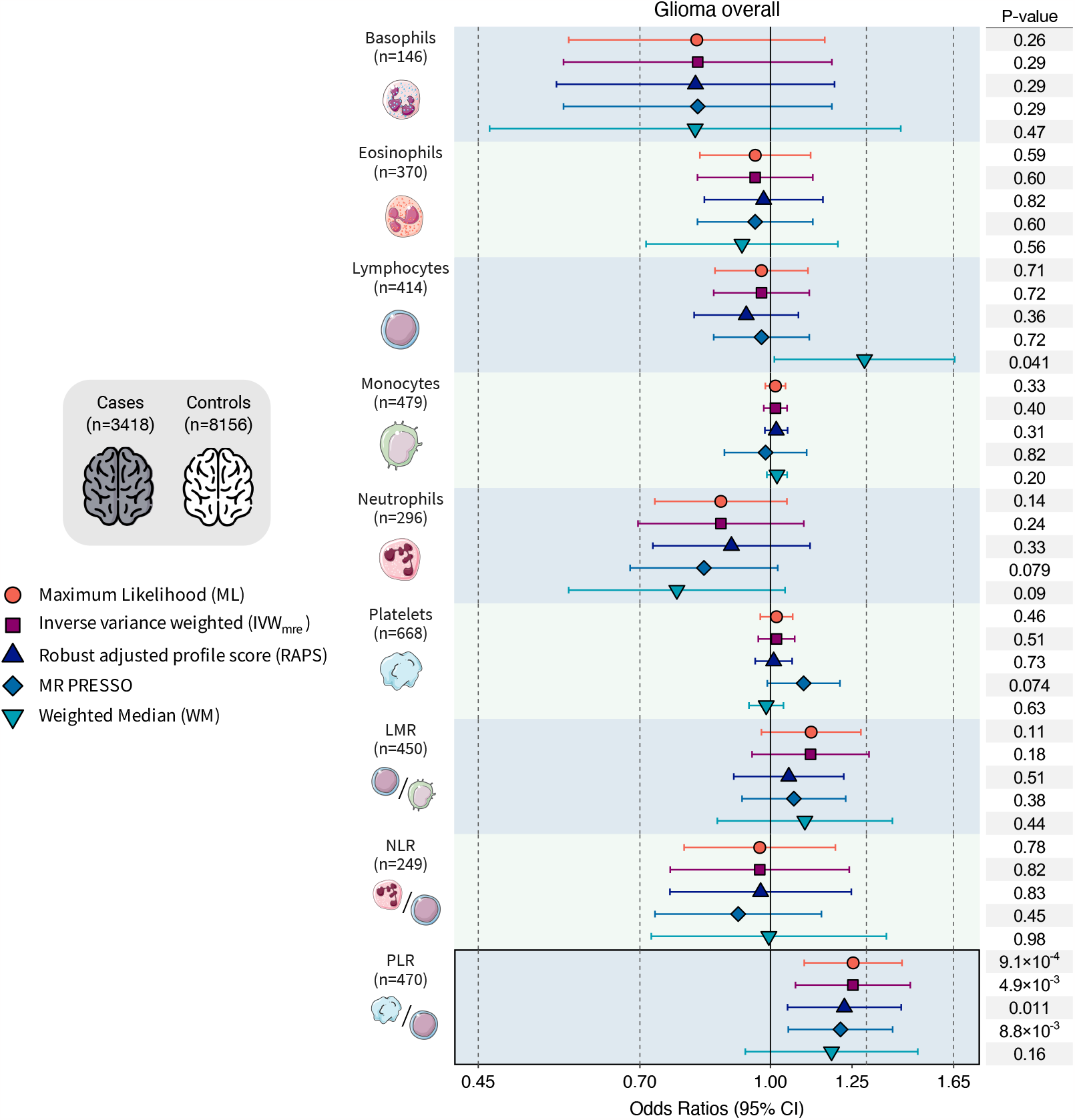
Mendelian randomization results for glioma overall. Visualization of odds ratios (OR) and 95% confidence intervals (CI) for the effect of increasing blood cell counts or blood cell ratios on the risk of glioma overall. For each blood cell phenotype, association results are shown for five Mendelian randomization estimation methods.

Genetically predicted blood cell traits differed for the major molecular subtypes of glioma. Genetic instruments for circulating blood cell profiles were primarily associated with susceptibility to IDH_mut_ gliomas, particularly IDH_mut-noncodel_ **(Figure 2; Supplementary Table S3)**, but not with IDH_wt_ gliomas, despite their larger sample size **(Figure 3; Supplementary Table S4)**. For most blood cell traits, Cochran’s Q (*P*_Q_<0.05) and the PRESSO Global test (*P*_Global_<0.05) detected heterogeneity among the causal effects implied by individual instruments, indicative of non-directional horizontal pleiotropy **(Supplementary Table S2)**; therefore, only associations that were statistically significant using pleiotropy-robust MR estimators were considered reliable.

**Figure 2:**
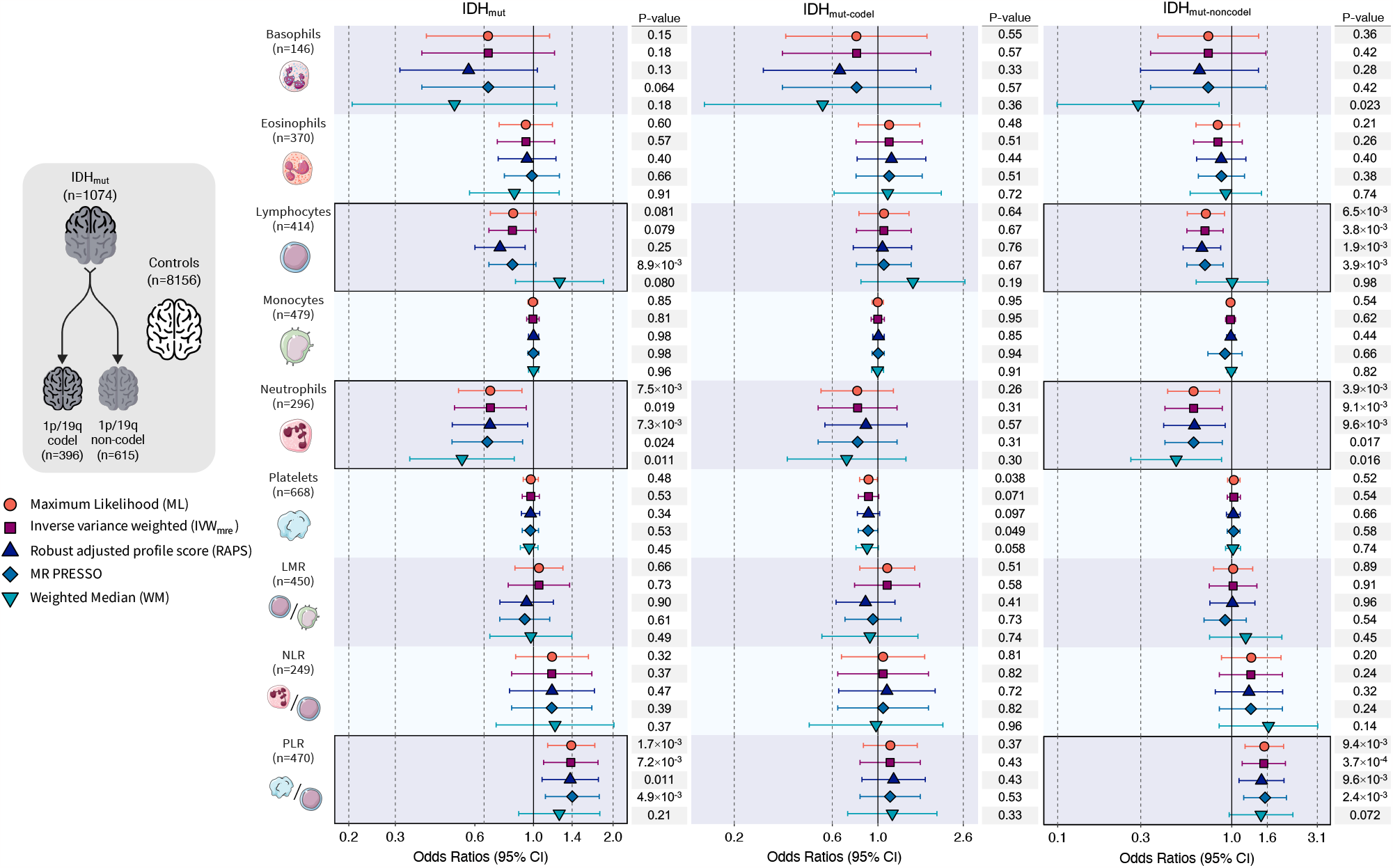
Mendelian randomization results for IDH mutated (IDH_mut_) glioma. Visualization of odds ratios (OR) and 95% confidence intervals (CI) for the effect of increasing blood cell counts or blood cell ratios on the risk of IDH_mut_ glioma and further stratified by the presence of 1p/19q co-deletion. For each blood cell phenotype, association results are shown for five Mendelian randomization estimation methods.

**Figure 3:**
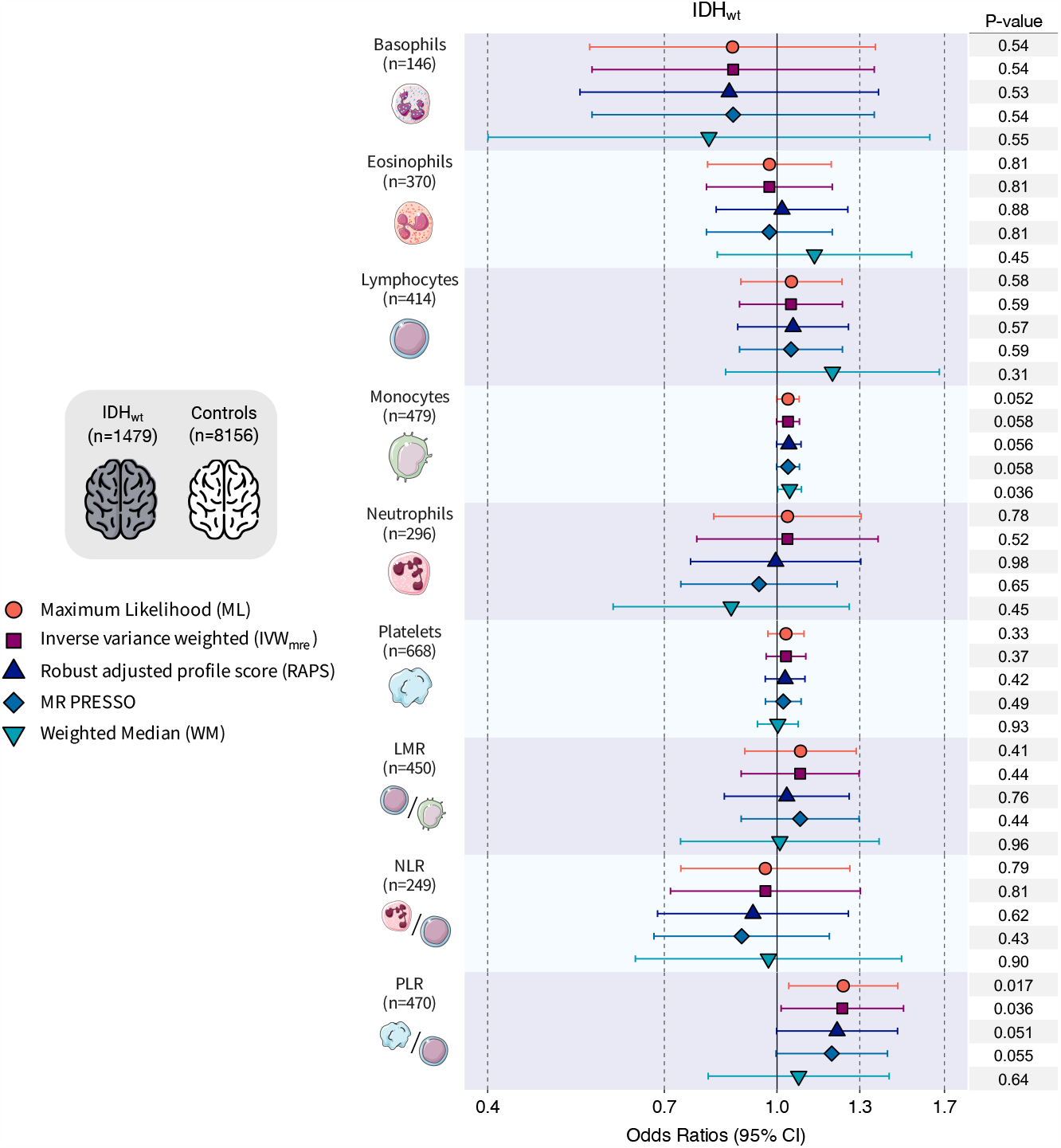
Mendelian randomization results for IDH wildtype (IDH_wt_) glioma. Visualization of Mendelian randomization odds ratios (OR) and 95% confidence intervals (CI) for the effect of increasing blood cell counts or blood cell ratios on the risk of IDH_wt_ glioma overall. For each blood cell phenotype, association results are shown for five Mendelian randomization estimation methods.

An increase in neutrophil counts was inversely associated with risk of IDH_mut_ tumors (OR_IVW-mre_=0.69, 0.50-0.94, p=0.019) and this was more pronounced for IDH_mut-nondel_ (OR_IVW-mre_=0.60, 0.41-0.88, p=9.1×10^−3^) subtypes **(Figure 2; Supplementary Table S3)**. Estimates were of the same magnitude in pleiotropy-corrected analyses for IDH_mut_ (OR_PRESSO_=0.67, p=0.011; OR_RAPS_=0.68, p=0.024) and IDH_mut-nondel_ (OR_PRESSO_=0.60, p=9.1×10^−3^; OR_RAPS_=0.61, p=0.017). Neutrophil counts were not significantly associated with risk of IDH_mut-codel_ tumors (OR_mre_=0.80, p=0.31), but the direction of effect was consistent with IDH_mut-nondel_. We also detected an inverse association between genetic predisposition to higher lymphocyte counts and risk of IDH_mut-nondel_ (OR_IVW-MRE_=0.70, 0.55-0.89, p=3.8×10^−3^).

The PLR association detected for glioma overall was also observed for IDH_mut_ (OR_IVW-MRE_=1.38, p=7.2×10^−3^), with a larger magnitude of effect among IDH_mut-noncodel_ (OR_ML_=1.54, p=9.4×10^−4^; OR_IVW-MRE_=1.53, p=3.7×10^−3^) than IDH_mut-codel_ tumors (OR_ML_=1.15, p=0.37) **(Supplementary Table S4)**. PLR was modestly associated with risk of IDH_wt_ tumors (OR_ML_=1.23, p=0.017), but this effect was reversed after correcting for directional pleiotropy (OR_Egger_=0.83, p=0.42). MR Egger analyses were not vulnerable to regression dilution bias due to measurement error^39^, as indicated by *I*^2^_GX_>0.98 for all phenotypes **(Supplementary Table S2)**. In addition to analytic correction for potentially invalid instruments we also manually removed them and re-fit MR IVW models **(Supplementary Table S5)**. These sensitivity analyses confirmed the subtype-specific associations observed for lymphocytes (IDH_mut_: OR=0.68, p=1.8×10^−4^; IDH_mut-noncodel_: OR=0.69, p=4.5×10^−4^), neutrophils (IDH_mut_: OR=0.65, p=1.4×10^−3^; IDH_mut-noncodel_: OR=0.61, p=5.2×10^−3^), and PLR (glioma: OR=1.27, p=2.2×10^−4^; IDH_mut_: OR=1.45, p=3.1×10^−4^; IDH_mut-noncodel_: OR=1.53, p=8.1×10^−4^).

Multivariable MR analyses suggested that only genetically predicted lymphocytes were independently associated with IDH_mut-noncodel_ glioma risk (OR_MV-IVW_=0.70, 0.48-1.00, p=0.051) **(Supplementary Table S6)**. Applying MV-LASSO to lymphocytes, neutrophils, platelets, and PLR resulted in the removal of platelets and shrunk the effect sizes of the remaining three traits (OR_MV-LASSO_=1.16 for PLR; OR_MV-LASSO_=0.89 for lymphocytes; OR_MV-LASSO_=0.90 for neutrophils). Only PLR (OR_MV-LASSO_=1.11) and neutrophils (OR_MV-LASSO_=0.86) were retained for IDH_mut_.

We also examined transcriptomic properties of the blood cell instruments for glioma-associated traits. Over 36% of variants (n=1014) were expression quantitative trait loci (eQTL) at FDR<0.05 in at least one of 13 brain tissues in GTEx v8 **(Figure S2)**. The prevalence of brain eQTLs ranged from 32% among instruments for platelets to 40% among instruments for basophils. Several target eGenes have been previously implicated in glioma susceptibility. Genetic instruments for eosinophils included eQTLs for *JAK1* and *D2HGDH*, neutrophil-associated eGenes included *NKD2* and *HEATR3*, and eQTLs for *STMN3, LIME1, RETL1*, and *GMEB2* were identified among instruments for platelets.

### Colocalization of glioma risk loci with blood cell traits

Among genetic instruments for blood cell traits, we identified 8 variants that reached genome-wide significance for at least one glioma phenotype **(Supplementary Table S7)**. Several of these variants were localized in regions known to be associated with telomere length, such as 5p15.33 (*TERT*) and 20q13.33 (*STMN3*-*RTEL1*). Since telomere maintenance is a known risk factor for glioma, it was incorporated into the fine-mapping and colocalization analyses. We detected evidence of a shared genetic signal at 2q37.3 (*D2HGDH*) for IDH_mut-nondel_ and eosinophil counts (PP=0.975), with evidence for rs34290285 as the causal variant (PP_SNP_=0.944) and an eQTL for *D2HGDH* in all 13 brain tissues. **(Supplementary Figure S3; Supplementary Table S8)**. The susceptibility signal for glioma overall and IDH_wt_ in 5p15.33 did not colocalize with NLR, but there was evidence of colocalization for IDH_wt_ with PLR, platelets, neutrophils, and telomere length (PP=0.994; rs7705526 PP_SNP_=1.0). The fine-mapped causal signals for IDH_mut_ in 8q24.21 appeared to be distinct from blood cell traits (PP=3.7×10^−38^). Lastly, in 20q13.33 there was strong evidence of colocalization for IDH_wt_ with platelets (PP=0.952), with weak support for rs6011018 as the causal variant (PP_SNP_=0.177). There was no evidence of colocalization with telomere length in this region.

### Prognostic associations for genetic predictors of blood cell traits

We explored whether genetic determinants of blood cell trait variation may predict disease outcomes in glioma patients. Survival analyses of each blood cell trait polygenic score (PGS) were conducted separately in each study **(Supplementary Table S9)** and meta-analyzed across the three case-series **(Figure 4; Supplementary Table S10)**. The meta-analysis identified four statistically significant (FDR<0.05) associations, all of which were restricted to the IDH_mut-codel_ subtype with 387 cases and 77 deaths. We observed an increased risk of mortality per SD increase in the PGS for basophils (HR=1.42, 95% CI: 1.08-1.86, p=0.012), eosinophils (HR=1.59, 1.18-2.14, p=2.4×10^−3^), lymphocytes (HR=1.65, 1.24-2.20, p=6.4×10^−4^), and neutrophils (HR=1.49, 1.13-1.97, p=5.2×10^−3^). There was evidence of heterogeneity for the neutrophil association (Cochran’s Q p-value=0.029, Higgins *I*^2^=0.79). Although the study-specific estimates had concordant directions of effect, the magnitude of the association was much larger in TCGA cases (HR=4.82, 1.63-14.29, p=4.5×10^−3^) than in the combined UCSF AGS and Mayo Clinic patient population (HR=1.37, 1.03-1.83, p=0.033).

**Figure 4:**
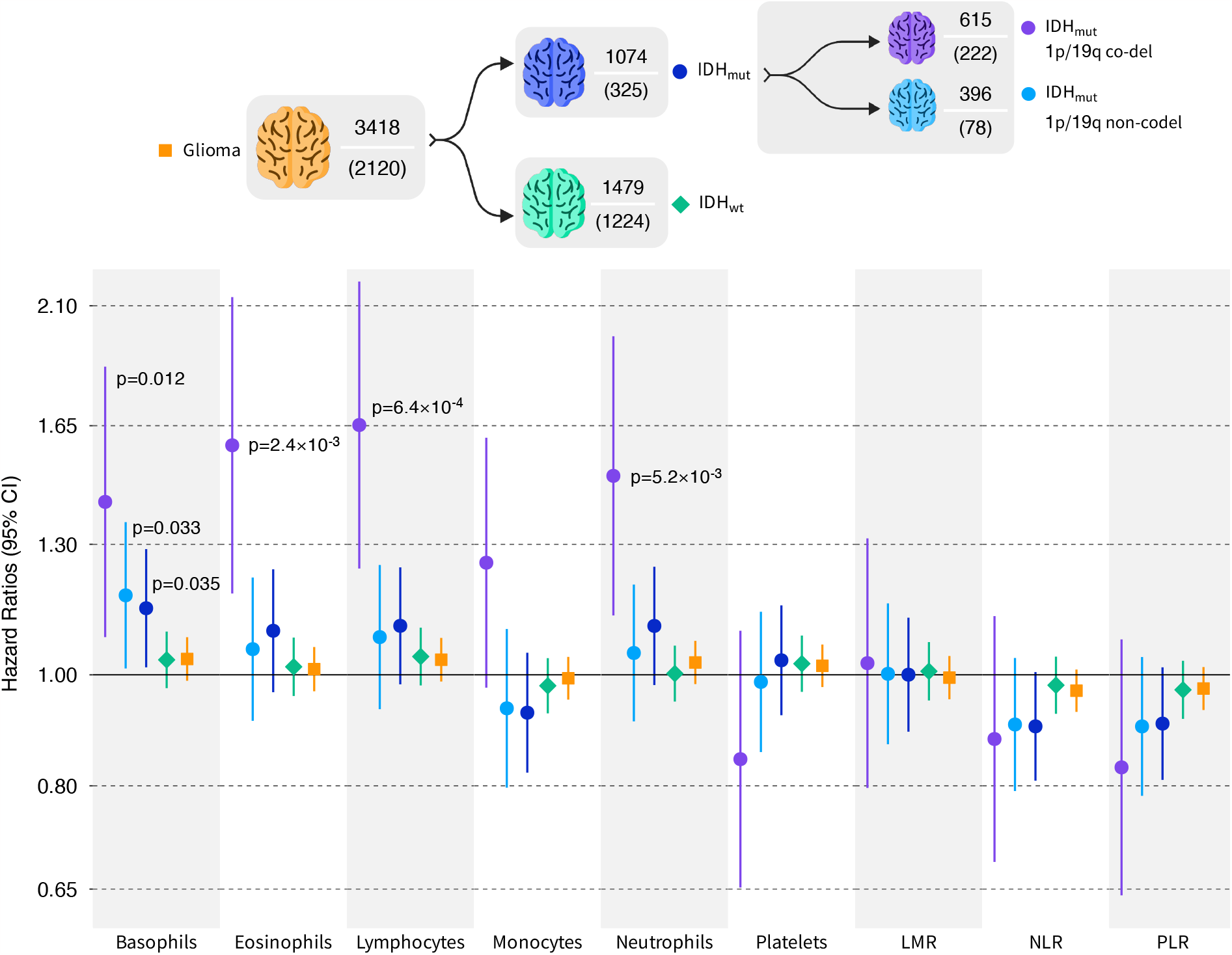
Meta-analysis of blood cell trait polygenic scores (PGS) associations with survival. Hazard ratios (HR) and 95% confidence intervals (CI) correspond to the effect of a standard deviation increase in the standardized PGS on all-cause mortality. P-values are reported for statistically significant associations. The sample size and corresponding number of mortality events is reported for each glioma subgroup.

Prior to evaluating associations with survival using Mendelian randomization, we tested for the presence of index event bias. Based on a set of pruned independent variants, the regression of survival log(HR) on incidence log(OR), with SIMEX adjustment for regression dilution, yielded a coefficient of −0.084 (95% CI: −0.101 to −0.072) for glioma and −0.137 (95% CI: −0.325 to −0.087) for IDH_mut_. This implies there are common causes of incidence and prognosis, the net effect of which produces modestly biased effects on survival for blood cell traits that are also associated with risk.

We present MR results based on unadjusted and bias-corrected instrument effects on survival, although the correction did not have an appreciable impact **(Supplementary Table S11)**. MR findings were generally concordant with PGS analyses, indicating an increased risk of death conferred by genetic predisposition to elevated counts of basophils (HR_IVW-MRE_=13.18, p=0.018), eosinophils (HR_IVW-MRE_=3.87, p=1.9×10^−3^), lymphocytes (HR_IVW-MRE_=4.36, p=3.2×10^−5^), and neutrophils (HR_IVW-MRE_=4.51, p=3.2×10^−3^). MR estimates were substantially larger than in PGS analyses, but they also had extremely wide 95% confidence intervals due to the large variance of SNP-specific effect sizes. Therefore, while we establish the direction of the putative causal effect of blood cell traits on glioma mortality, it is difficult to accurately infer its magnitude.

### Genetic associations with glioma tumor immune microenvironment features

To gain insight into potential tumorigenic mechanisms underlying the associations observed for blood cell traits, we examined their PGS associations with features of the heritable tumor immune microenvironment (TIME). Using data on 28 heritable TIME features identified by Sayaman et al.^38^ in TCGA, we conducted PGS analyses stratified by IDH mutation status **(Figure 5; Supplementary Table S12)**. No associations reached false discovery rate (FDR)<0.05 for IDH_mut_, but the neutrophil PGS was suggestively (FDR<0.10) associated with several signatures in the T-cell/Cytotoxic module, including enrichment scores for neutrophils, effector memory T-cells (T_EM_), and T-helper 17 cells (Th17). Among IDH_wt_ tumors the lymphocyte PGS was positively associated with CD8+ T-cell enrichment (p=1.3×10^−3^, FDR=0.036).

**Figure 5:**
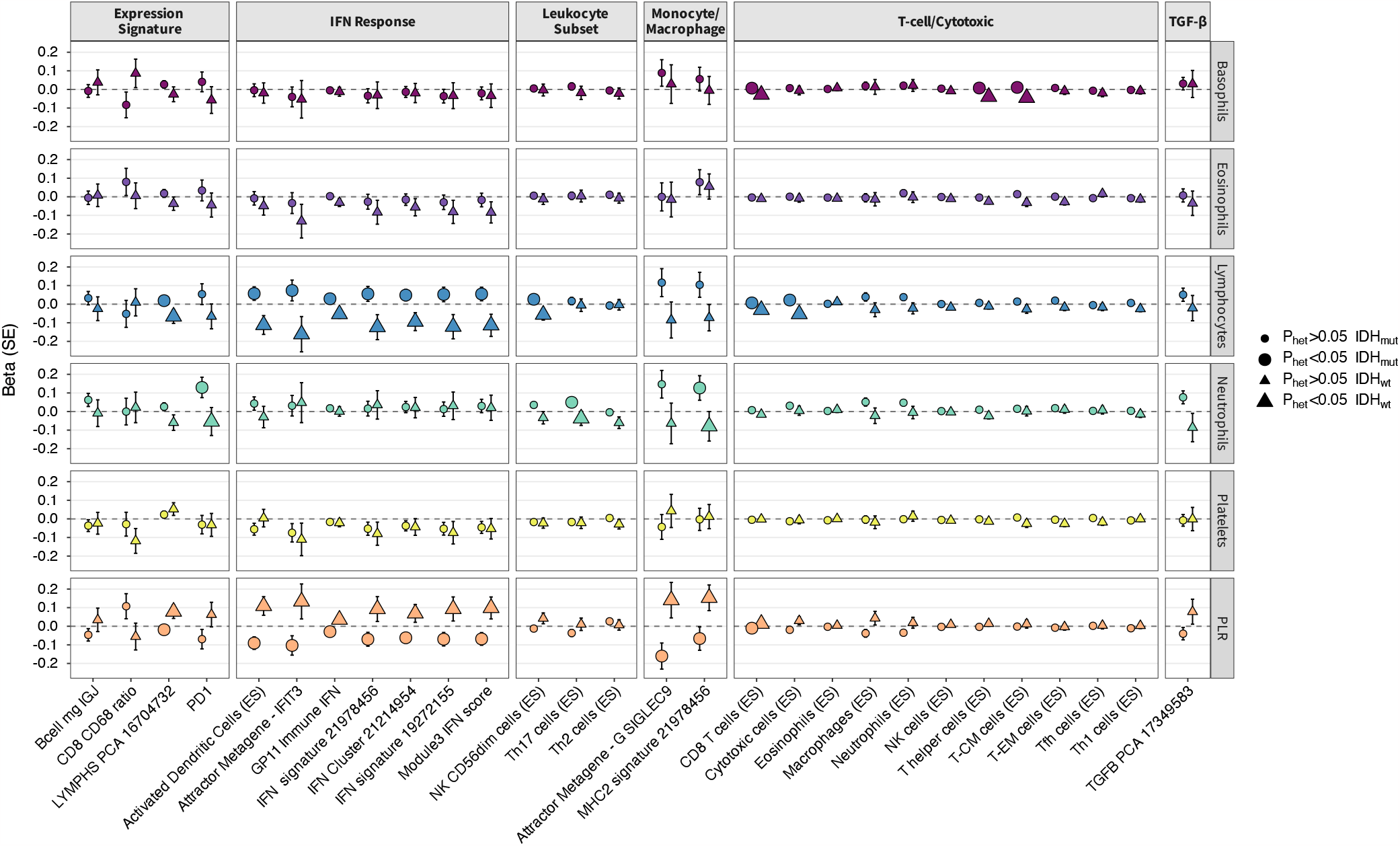
Associations of blood cell trait polygenic scores (PGS) with tumor immune microenvironment (TIME) features in TCGA. Associations for selected blood cell trait PGS with heritable tumor immune microenvironment features identified in TCGA by Sayaman et al. (2021). Heterogeneity in PGS effect on specific TIME features between IDH-mutated and IDH-wildtype tumors was tested using Cochran’s Q test. Beta coefficients for Enrichment Score (ES) phenotypes were estimated per 10-unit change.

There was significant effect size heterogeneity (*P*_het_<0.05) in the PGS associations for 17 traits when comparing estimates in IDH_mut_ and IDH_wt_ groups **(Figure 5)**. Most of the differential PGS effects were observed for TIME traits in IFN response and T-cell/Cytotoxic modules. Genetic scores for PLR and lymphocytes had the largest number of heterogeneous TIME associations (11 traits), followed by neutrophils and basophils (3 traits). The largest differences in genetic associations between IDH_mut_ and IDH_wt_ tumors were observed for activated dendritic cells (*P*_het_=8.9×10^−4^), CD8 T-cells (*P*_het_=8.2×10^−4^), T-helper cells (*P*_het_=4.3×10^−9^), multiple IFN signatures, and gene-based signatures G SIGLEC9 (*P*_het_=0.012), MHC II (*P*_het_=0.019), and PD-1 (*P*_het_=0.049).

## DISCUSSION

Blood cell counts are routinely measured in clinical settings to assess overall health, including systemic response to infection, inflammation, and allergic reactions. Additionally, blood cell counts are used during the course of cancer treatment to monitor adverse effects of chemotherapy and radiation. Markers of immune function, including blood cell proportions, have been associated with glioma survival^5,21^. However, their roles as etiologic factors have not been comprehensively investigated, partly because glioma and associated treatments influence immune cell profiles. We observed that genetic effects on disease risk and survival varied by glioma IDH mutation and 1p/19q co-deletion status. The heterogeneity in the effects of white blood cell counts and ratios across molecular subtypes of glioma suggests germline differences in susceptibility to immune-mediated risk factors. This complements previously observed differences in genetic susceptibility loci for these molecular subtypes^3,26^.

We found that genetic predisposition to higher circulating levels of lymphocytes and neutrophils were inversely associated with risk of IDH_mut_ gliomas, particularly the IDH_mut_ 1p/19q non-codeleted subtype. Colocalization of GWAS-identified glioma risk variants at 2q37.3 and 5p15.33 with blood cell traits also supports the existence of shared genetic mechanisms. The 2q37.3 (*D2HGDH*) risk locus for IDH_mut_ was discovered in the UCSF and Mayo Clinic study^26^ and the colocalized signal mapped to rs34290285, an instrument for eosinophils that has been linked to asthma and allergic diseases^40^. The protective effects observed for lymphocytes is broadly compatible with the hypothesis that this is a marker of improved immunosurveillance of cancer cells^41^. Genetic instruments in our study were developed in individuals without autoimmune or inflammatory conditions^22^, suggesting that benign elevation of lymphocytes may confer some advantages, such as improved recognition of precursor lesions or more efficient elimination of early cancers^41-43^.

Early versions of the immunosurveillance model attributed protective tumor immunity to antigen-specific lymphocytes, and it is now recognized that CD8+ and CD4+ T-cell responses play a critical role in immune clearance of tumor cells ^41,42^. Our genetic instruments lacked resolution at the cell-type level to investigate this directly, but our findings for lymphocytes are aligned with Ostrom et al.^17^, who reported a negative genetic correlation for lymphocyte percentage and risk of non-glioblastoma tumors, which are characterized by a high prevalence of IDH mutations. This study also detected negative genetic correlations with several autoimmune conditions and heritability enrichments among T cells, NK cells, and myeloid cells^17^, suggesting that immune activation modifies risk of glioma.

In contrast to lymphocytes, persistent neutrophil activation and infiltration is a hallmark of chronic inflammation associated with immunosuppression, tumor progression, and poor survival^44^. Neutrophils are the most abundant immune cell type, accounting for 50-70% of circulating leukocytes, and are the first line of defense against invading pathogens^44^. They have diverse functions and bridge between innate and adaptive immune response. Neutrophils are ascribed different roles in cancer depending on disease stage and can transition from exerting antitumor effects, such as CD8+ and CD4+ T cell priming, to cancer-promoting effects^44-46^. Circulating neutrophils are comprised of diverse cell populations displaying phenotypic heterogeneity and plasticity^44,47^. In addition to classic antitumorigenic (N1) and pro-tumorigenic (N2) neutrophils, there is also a distinction between high-density neutrophils (HDNs), characterized by an immunostimulatory N1-like profile, and low-density neutrophils (LDNs) that consist of immature myeloid-derived suppressor cells with an immunosuppressive phenotype^44,47^. Peripheral blood in healthy individuals predominantly consists of HDNs, while LDNs accumulate during chronic inflammation and malignancy^44,47^. Transforming growth factor β (TGF-β) can induce a transition from the HDN to the LDN phenotype, which is accompanied by loss of cytotoxic properties and gain of CD8+ T-cell suppression^44,47^. Although our genetic instruments do not distinguish neutrophil classes, the risk reducing effects observed for *IDH* mutated tumors may be reflect tumor suppressive properties associated with higher circulating levels of healthy neutrophil cell populations. This is also supported by in-vitro evidence that human polymorphonuclear neutrophils display innate abilities to target and destroy cancer cells^48^.

Our MR analysis uncovered that increased PLR, denoting a shift towards higher relative abundance of platelets to lymphocytes, conferred an increased risk of IDH_mut_ glioma, with the largest effect observed for IDH_mut_ tumors without 1p/19q non-codeletion. Although there was an attenuated, nominally significant PLR association for all gliomas combined, MR estimates for IDH_wt_ became reversed in sensitivity analyses, suggesting that the impact of PLR on disease susceptibility is likely restricted to IDH_mut_. Platelets patrol the circulatory system and become activated in response to vessel damage and bacterial infection. Wound-healing and proangiogenic properties of platelets, including stimulation of vascular endothelial growth factor (VEGF) and fibroblast growth factor, have been proposed as a plausible mechanism by which elevated platelet counts may predispose to tumor development^49,50^. However, platelet elevation alone was not associated with increased disease risk, suggesting that the mechanisms underlying the PLR association likely involve dysregulation of platelet interaction with lymphocytes.

Immune-sensing platelets participate in innate and adaptive responses, coordinate pro-inflammatory signaling, providing multiple candidate mechanisms through which platelets may contribute to carcinogenesis^49^. Platelets interact with lymphocytes through direct cell-to-cell contact and soluble signaling. Outside of the context of infection, interactions of platelets with immune cells have been shown to be involved in atherosclerosis and inflammation^51^, and emerging evidence suggests that platelets may dampen lymphocyte effector functions in the tumor microenvironment^50^. Platelets contain high levels of nonsignaling TGF-β and are the only cell population to constitutively express its docking receptor, glycoprotein A repetitions predominant (GARP), required for TGF-β activation^52,53^. In a murine model, TGF-β activated by platelet-expressed GARP suppressed T cell function and adoptive T cell therapy^53^. However, the interplay between systemic factors proxied by our genetic instruments and local immune response in the tumor is not well characterized^43^.

Our MR analysis did not implicate genetic regulation of blood cell traits as a risk factor for IDH_wt_ tumors. However, there was evidence of colocalization in 5p15.33 for rs7705526, an IDH_wt_ risk variant, with neutrophils, platelets, PLR, and telomere length. This is consistent with previous work showing multiple candidate mechanisms in this highly pleiotropic locus, not limited to telomere maintenance^54^. There was no evidence of colocalization in 8q24, consistent with recent work demonstrating that the causal risk variant at this locus exerts its effects through interaction with the *Myc* promoter and increased *Myc* expression^55^.

Our survival analyses identified statistically significant associations with polygenic scores for blood cell traits only in IDH_mut_ 1p/19q co-deleted tumors, which may partly reflect longer median survival in these patients and greater power to detect factors that modify survival trajectories. In contrast to disease susceptibility, genetic predisposition to higher circulating counts of basophils, eosinophils, lymphocytes, and neutrophils were associated with higher risk of early death. Associations with survival in the same direction, albeit non-significant, were detected across IDH_mut_ gliomas, yet we observe no evidence for an association in IDH_wt_ tumors. These findings may reflect context-dependent effects, especially for neutrophils, but we also note that opposing effects on disease risk and survival were not detected within the same tumor subtype – MR risk estimates for the effect of blood cell traits on IDH_mut_ 1p/19q co-deleted tumors were null. The impact of genetic predictors of blood cell traits on survival may interact with somatic mutations within the tumor resulting in differences in TIME. This is supported by heterogeneity in blood cell PGS associations with TME features in TCGA glioma cases, stratified by IDH mutation status.

Differences in the immune landscape of IDH_mut_ and IDH_wt_ tumors are well established and attributed to the accumulation of the D-2HG oncometabolite produced by the mutant IDH enzyme, which results in widespread hypermethylation and gene repression^56^. In addition to downregulating immune-related signaling, D-2HG has been shown to directly suppress T cell activation and lower the expression MHC class II, CD80, and CD86 molecules, further reducing capacity for antigen presentation^56,57^. This aligns with our observations of significant heterogeneity in basophil, neutrophil, and lymphocyte PGS effects on MHC-II, CD8 T cell, and cytotoxic T cell TIME signatures.

The neutrophil PGS was nominally associated with programmed cell death protein-1 (PD-1) TIME signature in IDH_mut_ tumors only and this PGS effect showed significant heterogeneity by IDH status. PD-1 and its ligand PD-L1 have been shown to delay neutrophil apoptosis^58^, which may be related to our inverse risk association with neutrophils in IDH_mut_ tumors. PD-1/PD-L1 check point inhibitors have been of interest as a potential glioma therapeutic, however most studies have focused on glioblastoma tumors (IDH_wt_ according to WHO 2021) with limited success^59^. Our results suggest that therapeutic intervention using anti-PD-1/PD-L1 approaches may vary by IDH status and germline predisposition. Considering the impact of genetically driven changes in immune cell types on survival may refine the selection of patients into clinical trials or help inform more personalized management strategies. We also observed differences in associations across interferon responses and PGS for lymphocytes between IDH_wt_ and IDH_mut_ tumors. Interferons such as IFN-γ can directly affect lymphocyte function, including cytotoxic CD8+ T-cells. Recent studies have highlighted the role of IFN-γ in glioblastoma, showing that reducing Notch-regulated IFN-γ signaling aids in the evasion of immune surveillance and reduces tumor clearance^60^. IDH_mut-codel_ gliomas maintain the same immunologically cold phenotype as overall IDH_mut_ gliomas, but little is known about specific effects of 1p/19q co-deletion on the TIME^61^. As IDH mutations alone are not sufficient for gliomagenesis^62^, interactions with additional somatic events, potentially the 1p/19 co-deletion, may contribute to the observed results.

Taken together, our study builds on decades of research that has implicated aspects of immune function in the etiology of glioma. While findings of observational studies have been heterogeneous and vulnerable to reporting biases and reverse causation, our study provides robust evidence, as genetic predictors are not affected by external factors that have disruptive effects on immune cell distributions, such as infections and treatment mediated effects. Our analysis benefits from the strength of the genetic instruments, which explain between 4.5% and 23.5% of target trait variation^22^, and range of statistical approaches for evaluating the robustness of our conclusions to genetic confounding by pleiotropy. Although we provide statistical evidence that variation in immune cell counts may influence risk of IDH_mut_ glioma, mechanistic studies will be required to validate this relationship and elucidate relevant cell types and processes. We refined our MR and survival results by molecular subtypes of glioma, which sets our work apart from other studies and enhances its clinical relevance. However, a caveat for such analyses is the loss of statistical power. Future studies with larger sample sizes will be required to replicate these findings and consider additional tumor features, such as *TERT* and *ATRX* mutations. As the search for the causes of glioma and improved treatments of this highly complex and deadly cancer continues, interrogating the genetic architecture of disease susceptibility and progression will contribute valuable insights.

## Data Availability

Genotype data of control samples from the 1958 British Birth Cohort and UK Blood Service Control Group were made available from the Wellcome Trust Case Control Consortium (WTCCC) and downloaded from the European Genotype Archive under ascension numbers EGAD00000000021 and EGAD00000000023, respectively. Genotype data of glioma cases from The Cancer Genome Atlas (TCGA) were obtained from Database of Genotypes and Phenotypes (dbGaP) (phs000178). Genotype data of control samples from the Glioma International Case Control Study (GICC) are available from dbGaP under accession phs001319.v1.p1.

## ACKNOWLEDGEMENTS

LK is supported by funding from the National Institutes of Health (NIH): R00CA246076. Work at University of California, San Francisco was supported by the NIH (grant numbers T32CA151022, R01CA52689, P50CA097257, R01CA126831, R01CA139020, R01AI128775, and R25CA112355), the National Brain Tumor Foundation, the Stanley D. Lewis and Virginia S. Lewis Endowed Chair in Brain Tumor Research, the Robert Magnin Newman Endowed Chair in Neuro-oncology, and by donations from families and friends of John Berardi, Helen Glaser, Elvera Olsen, Raymond E. Cooper, and William Martinusen. This publication was supported by the National Center for Research Resources and the National Center for Advancing Translational Sciences, National Institutes of Health, through UCSF-CTSI grant number UL1 RR024131. Its contents are solely the responsibility of the authors and do not necessarily represent the official views of the NIH. The collection of cancer incidence data used in this study was supported by the California Department of Public Health pursuant to California Health and Safety Code Section 103885; Centers for Disease Control and Prevention’s (CDC) National Program of Cancer Registries, under cooperative agreement 5NU58DP006344; the National Cancer Institute’s Surveillance, Epidemiology, and End Results Program under contract HHSN261201800032I awarded to the University of California, San Francisco, contract HHSN261201800015I awarded to the University of Southern California, and contract HHSN261201800009I awarded to the Public Health Institute, Cancer Registry of Greater California. The ideas and opinions expressed herein are those of the author(s) and do not necessarily reflect the opinions of the State of California, Department of Public Health, the National Cancer Institute, and the Centers for Disease Control and Prevention or their contractors and subcontractors. Work at Mayo was supported by NIH grants CA230712, P50 CA108961, and CA139020; the National Brain Tumor Society; the loglio Collective; the Mayo Clinic; and the Ting Tsung and Wei Fong Chao Foundation.

