## Supplementary Tables S1-S12, Supplementary Figures S1-S3 for "Genetic predisposition to altered blood cell homeostasis is associated with glioma risk and survival"

**Supplementary Table S1: Mendelian randomization results for glioma overall.** Odds ratios (OR) and 95% confidence intervals (CI) for the effect of increasing blood cell counts or cell type ratios on the risk of glioma. MR Egger results are only presented for blood cell traits with statistically significant evidence of directional pleiotropy (MR Egger intercept $\beta_0 \neq 0$, $p<0.05$).

| Blood Cell Trait | MR Estimator | $N_{SNP}$ | Explained Variance | Glioma Overall | | |
|---|---|---|---|---|---|---|
| | | | | OR | (95% CI) | P |
| Basophils | IVW-MRE | 146 | 0.045 | 0.82 | (0.57 – 1.18) | 0.29 |
| | Maximum likelihood | | | 0.82 | (0.58 – 1.16) | 0.26 |
| | Weighted median | | | 0.81 | (0.46 – 1.43) | 0.47 |
| | RAPS | | | 0.81 | (0.56 – 1.19) | 0.29 |
| | PRESSO | | | 0.82 | (0.57 – 1.18) | 0.29 |
| Eosinophils | IVW-MRE | 370 | 0.105 | 0.96 | (0.82 – 1.12) | 0.60 |
| | Maximum likelihood | | | 0.96 | (0.82 – 1.12) | 0.59 |
| | Weighted median | | | 0.93 | (0.71 – 1.20) | 0.56 |
| | RAPS | | | 0.98 | (0.83 – 1.15) | 0.82 |
| | MR Egger | | | 0.67 | (0.46 – 0.99) | 0.044 |
| | PRESSO | | | 0.96 | (0.82 – 1.12) | 0.60 |
| Lymphocytes | IVW-MRE | 414 | 0.114 | 0.98 | (0.86 – 1.11) | 0.72 |
| | Maximum likelihood | | | 0.98 | (0.86 – 1.11) | 0.71 |
| | Weighted median | | | 1.29 | (1.01 – 1.65) | 0.041 |
| | RAPS | | | 0.94 | (0.81 – 1.08) | 0.36 |
| | MR Egger | | | 1.23 | (0.98 – 1.54) | 0.075 |
| | PRESSO | | | 0.98 | (0.89 – 1.11) | 0.72 |
| Monocytes | IVW-MRE | 479 | 0.159 | 1.01 | (0.98 – 1.05) | 0.40 |
| | Maximum likelihood | | | 1.01 | (0.99 – 1.04) | 0.33 |
| | Weighted median | | | 1.02 | (0.99 – 1.05) | 0.20 |
| | RAPS | | | 1.02 | (0.99 – 1.05) | 0.31 |
| | MR Egger | | | 1.02 | (0.99 – 1.06) | 0.19 |
| | PRESSO | | | 0.99 | (0.88 – 1.10) | 0.82 |
| Neutrophils | IVW-MRE | 296 | 0.082 | 0.87 | (0.70 – 1.10) | 0.24 |
| | Maximum likelihood | | | 0.87 | (0.73 – 1.05) | 0.14 |
| | Weighted median | | | 0.77 | (0.58 – 1.04) | 0.090 |
| | RAPS | | | 0.90 | (0.73 – 1.11) | 0.33 |
| | PRESSO | | | 0.83 | (0.68 – 1.02) | 0.079 |
| Platelets | IVW-MRE | 668 | 0.235 | 1.02 | (0.97 – 1.07) | 0.51 |
| | Maximum likelihood | | | 1.02 | (0.97 – 1.06) | 0.46 |
| | Weighted median | | | 0.99 | (0.94 – 1.04) | 0.63 |
| | RAPS | | | 1.01 | (0.96 – 1.06) | 0.73 |
| | MR Egger | | | 0.99 | (0.94 – 1.05) | 0.76 |

| | | | | | | |
|---|---|---|---|---|---|---|
| | PRESSO | | | 1.10 | (0.99 – 1.21) | 0.074 |
| LMR | IVW-MRE | 450 | 0.147 | 1.12 | (0.95 – 1.31) | 0.18 |
| | Maximum likelihood | | | 1.12 | (0.98 – 1.28) | 0.11 |
| | Weighted median | | | 1.10 | (0.86 – 1.40) | 0.44 |
| | RAPS | | | 1.05 | (0.90 – 1.22) | 0.51 |
| | PRESSO | | | 1.07 | (0.93 – 1.23) | 0.38 |
| NLR | IVW-MRE | 249 | 0.063 | 0.97 | (0.76 – 1.24) | 0.82 |
| | Maximum likelihood | | | 0.97 | (0.79 – 1.19) | 0.78 |
| | Weighted median | | | 1.00 | (0.72 – 1.37) | 0.98 |
| | RAPS | | | 0.97 | (0.76 – 1.25) | 0.83 |
| | PRESSO | | | 0.92 | (0.73 – 1.15) | 0.45 |
| PLR | IVW-MRE | 470 | 0.145 | 1.25 | (1.07 – 1.47) | $4.9\times10^{-3}$ |
| | Maximum likelihood | | | 1.25 | (1.10 – 1.43) | $9.1\times10^{-4}$ |
| | Weighted median | | | 1.18 | (0.93 – 1.50) | 0.16 |
| | RAPS | | | 1.22 | (1.05 – 1.43) | 0.011 |
| | PRESSO | | | 1.21 | (1.05 – 1.40) | $8.8\times10^{-3}$ |

[1] Out-of-sample trait variance explained by the genetic instruments was previously reported in Kachuri et al. (2021)[22] and estimated for the subset of genetic instruments that were available for this analysis

Abbreviations:

| | |
|---|---|
| IVW-MRE | Inverse variance weighted multiplicative random effect |
| RAPS | Robust adjusted profile score |
| PRESSO | Pleiotropy Residual Sum and Outlier |
| LMR | Lymphocyte to monocyte ratio |
| NLR | Neutrophil to lymphocyte ratio |
| PLR | Platelet to lymphocyte ratio |

**Supplementary Table S2: Results of Mendelian Randomization diagnostic tests.** Assessment of the robustness of underlying Mendelian randomization assumptions for each set of analyses of glioma susceptibility, overall and stratified by major molecular subtypes.

| Blood Cell Trait | $I^2_{GX}$ | Glioma: Heterogeneity[1] | | | MR Egger[2] | | MR PRESSO | IDH-mutated ($IDH_{mut}$): Heterogeneity[1] | | | MR Egger[2] | | MR PRESSO |
|---|---|---|---|---|---|---|---|---|---|---|---|---|---|
| | | Q | DF | $P_{Q\text{-value}}$ | $\beta_0$ | $P_{Egger}$ | $P_{Global}$[3] | Q | DF | $P_{Q\text{-value}}$ | $\beta_0$ | $P_{Egger}$ | $P_{Global}$[3] |
| Basophils | 0.985 | 162.1 | 145 | 0.16 | -0.011 | 0.24 | 0.15 | 171.3 | 145 | 0.067 | -0.008 | 0.58 | 0.073 |
| Eosinophils | 0.986 | 406.2 | 369 | 0.089 | 0.012 | 0.048 | 0.091 | 432.7 | 369 | 0.012 | 0.006 | 0.51 | 0.011 |
| Lymphocytes | 0.986 | 464.3 | 413 | 0.041 | -0.010 | 0.015 | 0.031 | 484.7 | 413 | $8.5\times10^{-3}$ | -0.009 | 0.14 | $8.0\times10^{-3}$ |
| Monocytes | 0.988 | 639.9 | 478 | $9.6\times10^{-7}$ | -0.005 | 0.048 | 0.013 | 765.8 | 478 | $1.1\times10^{-15}$ | -0.006 | 0.19 | $2.5\times10^{-4}$ |
| Neutrophils | 0.985 | 476.1 | 295 | $1.1\times10^{-10}$ | 0.007 | 0.37 | $<1.0\times10^{-4}$ | 385.2 | 295 | $3.2\times10^{-4}$ | 0.013 | 0.23 | $3.0\times10^{-4}$ |
| Platelets | 0.988 | 906.6 | 667 | $1.6\times10^{-9}$ | 0.005 | 0.031 | $6.7\times10^{-5}$ | 858.6 | 667 | $6.8\times10^{-7}$ | 0.004 | 0.24 | $6.7\times10^{-5}$ |
| LMR | 0.988 | 631.9 | 449 | $2.5\times10^{-8}$ | -0.005 | 0.33 | $<6.7\times10^{-5}$ | 753.2 | 449 | $9.3\times10^{-18}$ | -0.007 | 0.42 | $<6.7\times10^{-5}$ |
| NLR | 0.985 | 355.0 | 248 | $9.5\times10^{-6}$ | 0.013 | 0.16 | $<1e\times10^{-4}$ | 306.7 | 248 | $6.5\times10^{-3}$ | -0.008 | 0.54 | $6.2\times10^{-3}$ |
| PLR | 0.987 | 658.8 | 469 | $1.5\times10^{-8}$ | 0.009 | 0.11 | $<6.7\times10^{-5}$ | 635.5 | 469 | $4.3\times10^{-7}$ | 0.009 | 0.30 | $<6.7\times10^{-5}$ |

| | $I^2_{GX}$ | IDH-mutated 1p/19q co-deleted ($IDH_{mut\text{-}codel}$): Heterogeneity[1] | | | MR Egger[2] | | MR PRESSO | IDH-mutated 1p/19q non-codeleted ($IDH_{mut\text{-}noncodel}$): Heterogeneity[1] | | | MR Egger[2] | | MR PRESSO |
|---|---|---|---|---|---|---|---|---|---|---|---|---|---|
| | | Q | DF | $P_{Q\text{-value}}$ | $\beta_0$ | $P_{Egger}$ | $P_{Global}$[3] | Q | DF | $P_{Q\text{-value}}$ | $\beta_0$ | $P_{Egger}$ | $P_{Global}$[3] |
| Basophils | 0.985 | 162.8 | 145 | 0.15 | -0.018 | 0.38 | 0.14 | 194.0 | 145 | $4.1\times10^{-3}$ | -0.010 | 0.58 | $4.3\times10^{-3}$ |
| Eosinophils | 0.986 | 436.8 | 369 | $8.6\times10^{-3}$ | -0.008 | 0.59 | $8.1\times10^{-3}$ | 468.8 | 369 | $3.2\times10^{-4}$ | 0.017 | 0.17 | $3.6\times10^{-4}$ |
| Lymphocytes | 0.986 | 502.4 | 413 | $1.7\times10^{-3}$ | -0.016 | 0.086 | $1.8\times10^{-3}$ | 430.1 | 413 | 0.27 | -0.007 | 0.34 | 0.26 |
| Monocytes | 0.988 | 665.9 | 478 | $2.6\times10^{-8}$ | -0.005 | 0.38 | $1.3\times10^{-3}$ | 716.8 | 478 | $8.0\times10^{-12}$ | -0.005 | 0.32 | $2.8\times10^{-3}$ |
| Neutrophils | 0.985 | 356.9 | 295 | $7.9\times10^{-3}$ | -0.006 | 0.71 | $7.7\times10^{-3}$ | 367.7 | 295 | $2.5\times10^{-3}$ | 0.018 | 0.19 | $2.4\times10^{-3}$ |
| Platelets | 0.988 | 931.4 | 667 | $4.7\times10^{-11}$ | 0.007 | 0.21 | $6.7\times10^{-5}$ | 752.9 | 667 | 0.011 | 0.004 | 0.28 | 0.019 |
| LMR | 0.988 | 645.2 | 449 | $3.3\times10^{-9}$ | 0.001 | 0.92 | $<6.7\times10^{-5}$ | 681.6 | 449 | $7.8\times10^{-12}$ | -0.014 | 0.19 | $<6.7\times10^{-5}$ |
| NLR | 0.985 | 300.1 | 248 | 0.013 | 0.008 | 0.66 | 0.012 | 283.4 | 248 | 0.061 | -0.020 | 0.19 | 0.055 |
| PLR | 0.987 | 596.8 | 469 | $5.5\times10^{-5}$ | 0.012 | 0.35 | $6.7\times10^{-5}$ | 600.0 | 469 | $3.8\times10^{-12}$ | 0.015 | 0.15 | $<6.7\times10^{-5}$ |

| | $I^2_{GX}$ | Heterogeneity[1] | | | MR Egger[2] | | MR PRESSO |
|---|---|---|---|---|---|---|---|
| | IDH wildtype ($IDH_{wt}$) | | | | | | |
| | | Q | DF | $P_{Q\text{-value}}$ | $\beta_0$ | $P_{Egger}$ | $P_{Global}$[3] |
| Basophils | 0.985 | 143.6 | 145 | 0.52 | -0.014 | 0.22 | 0.53 |
| Eosinophils | 0.986 | 387.6 | 369 | 0.24 | 0.006 | 0.45 | 0.24 |
| Lymphocytes | 0.986 | 431.3 | 413 | 0.26 | -0.006 | 0.36 | 0.26 |
| Monocytes | 0.988 | 506.5 | 478 | 0.18 | -0.009 | 0.082 | 0.32 |
| Neutrophils | 0.985 | 456.6 | 295 | $4.5\times10^{-9}$ | -0.005 | 0.084 | $<1.0\times10^{-4}$ |
| Platelets | 0.988 | 857.7 | 667 | $7.5\times10^{-7}$ | 0.000 | 0.98 | $1.3\times10^{-4}$ |
| LMR | 0.988 | 510.4 | 449 | 0.024 | 0.024 | 0.029 | 0.025 |
| NLR | 0.985 | 319.4 | 248 | $1.5\times10^{-3}$ | 0.003 | 0.26 | $1.9\times10^{-3}$ |
| PLR | 0.987 | 601.0 | 469 | $3.4\times10^{-5}$ | 0.014 | 0.059 | $<6.7\times10^{-5}$ |

[1] Modified Cochran's Q test for heterogeneity in causal effect estimates (indicative of horizontal pleiotropy), p-values<0.05 are considered statistically significant

[2] Tests for directional horizontal pleiotropy indicated by non-zero in MR Egger intercept ($\beta_0$) values, p-values<0.05 are considered statistically significant

[3] Tests for horizontal pleiotropy, empirical p-values were estimated based on 12000-15000 replicates, p-values<0.05 are considered statistically significant

[4] Values of $I^2_{GX}<0.90$ indicate potential for regression dilution bias and violation of the NOME (no measurement error assumption)

Abbreviations:

PRESSO Pleiotropy Residual Sum and Outlier
LMR Lymphocyte to monocyte ratio
NLR Neutrophil to lymphocyte ratio
PLR Platelet to lymphocyte ratio

**Supplementary Table S3: Mendelian randomization results for IDH mutant ($IDH_{mut}$) glioma.** Odds ratios (OR) and 95% confidence intervals (CI) estimate the effect of increasing blood cell counts or cell type ratios on disease risk. MR Egger results are only presented for traits with statistically significant evidence of directional pleiotropy (MR Egger intercept $\beta_0 \neq 0$, $p<0.05$).

| Blood Cell Trait | MR Estimator | $N_{SNP}$ | $IDH_{mut}$ | | | $IDH_{mut}$ 1p/19q co-deleted | | | $IDH_{mut}$ 1p/19q non-codeleted | | |
|---|---|---|---|---|---|---|---|---|---|---|---|
| | | | OR | (95% CI) | P | OR | (95% CI) | P | OR | (95% CI) | P |
| Basophils | IVW-MRE | 146 | 0.67 | (0.38 – 1.20) | 0.18 | 0.79 | (0.34 – 1.18) | 0.57 | 0.73 | (0.34 – 1.57) | 0.42 |
| | Maximum likelihood | | 0.67 | (0.39 – 1.15) | 0.15 | 0.79 | (0.36 – 1.73) | 0.55 | 0.73 | (0.38 – 1.43) | 0.36 |
| | Weighted median | | 0.50 | (0.21 – 1.22) | 0.13 | 0.54 | (0.14 – 2.02) | 0.36 | 0.29 | (0.10 – 0.85) | 0.023 |
| | RAPS | | 0.57 | (0.31 – 1.03) | 0.064 | 0.65 | (0.28 – 1.53) | 0.33 | 0.65 | (0.30 – 1.42) | 0.28 |
| | PRESSO | | 0.67 | (0.38 – 1.20) | 0.18 | 0.79 | (0.34 – 1.18) | 0.57 | 0.73 | (0.34 – 1.57) | 0.42 |
| Eosinophils | IVW-MRE | 370 | 0.94 | (0.73 – 1.20) | 0.60 | 1.13 | (0.78 – 1.64) | 0.51 | 0.83 | (0.60 – 1.15) | 0.26 |
| | Maximum likelihood | | 0.93 | (0.74 – 1.18) | 0.57 | 1.13 | (0.80 – 1.60) | 0.48 | 0.83 | (0.62 – 1.11) | 0.21 |
| | Weighted median | | 0.85 | (0.57 – 1.25) | 0.40 | 1.12 | (0.61 – 2.03) | 0.72 | 0.92 | (0.58 – 1.48) | 0.74 |
| | RAPS | | 0.95 | (0.73 – 1.22) | 0.66 | 1.16 | (0.79 – 1.71) | 0.44 | 0.87 | (0.63 – 1.21) | 0.40 |
| | PRESSO | | 0.99 | (0.78 – 1.25) | 0.91 | 1.13 | (0.78 – 1.64) | 0.51 | 0.87 | (0.64 – 1.18) | 0.38 |
| Lymphocytes | IVW-MRE | 414 | 0.83 | (0.68 – 1.02) | 0.079 | 1.07 | (0.79 – 1.45) | 0.67 | 0.70 | (0.55 – 0.89) | $3.8\times10^{-3}$ |
| | Maximum likelihood | | 0.84 | (0.69 – 1.02) | 0.081 | 1.07 | (0.81 – 1.42) | 0.64 | 0.71 | (0.55 – 0.91) | $6.5\times10^{-3}$ |
| | Weighted median | | 1.25 | (0.85 – 1.84) | 0.25 | 1.48 | (0.83 – 2.65) | 0.19 | 1.00 | (0.62 – 1.62) | 0.98 |
| | RAPS | | 0.75 | (0.60 – 0.93) | $8.9\times10^{-3}$ | 1.05 | (0.76 – 1.46) | 0.76 | 0.67 | (0.53 – 0.86) | $1.9\times10^{-3}$ |
| | PRESSO | | 0.83 | (0.68 – 1.02) | 0.080 | 1.07 | (0.79 – 1.45) | 0.67 | 0.70 | (0.55 – 0.89) | $3.9\times10^{-3}$ |
| Monocytes | IVW-MRE | 479 | 0.99 | (0.94 – 1.05) | 0.85 | 1.00 | (0.93 – 1.07) | 0.95 | 0.98 | (0.92 – 1.05) | 0.62 |
| | Maximum likelihood | | 0.99 | (0.95 – 1.04) | 0.81 | 1.00 | (0.94 – 1.06) | 0.95 | 0.98 | (0.93 – 1.04) | 0.54 |
| | Weighted median | | 1.00 | (0.96 – 1.04) | 0.98 | 1.00 | (0.93 – 1.06) | 0.91 | 0.99 | (0.94 – 1.05) | 0.82 |
| | RAPS | | 1.00 | (0.96 – 1.05) | 0.98 | 1.01 | (0.94 – 1.07) | 0.85 | 0.99 | (0.94 – 1.04) | 0.66 |
| | PRESSO | | 1.00 | (0.95 – 1.05) | 0.96 | 1.00 | (0.94 – 1.07) | 0.94 | 0.91 | (0.73 – 1.15) | 0.44 |
| Neutrophils | IVW-MRE | 296 | 0.69 | (0.50 – 0.94) | 0.019 | 0.80 | (0.51 – 1.24) | 0.31 | 0.60 | (0.41 – 0.88) | $9.1\times10^{-3}$ |
| | Maximum likelihood | | 0.69 | (0.52 – 0.90) | $7.5\times10^{-3}$ | 0.79 | (0.53 – 1.19) | 0.26 | 0.60 | (0.43 – 0.85) | $3.9\times10^{-3}$ |
| | Weighted median | | 0.54 | (0.34 – 0.85) | $7.3\times10^{-3}$ | 0.70 | (0.36 – 1.37) | 0.30 | 0.48 | (0.26 – 0.87) | 0.016 |
| | RAPS | | 0.68 | (0.49 – 0.95) | 0.024 | 0.87 | (0.55 – 1.38) | 0.57 | 0.61 | (0.41 – 0.92) | 0.017 |

| | | | | | | | | | | | |
|---|---|---|---|---|---|---|---|---|---|---|---|
| | PRESSO | | 0.67 | (0.49 – 0.91) | 0.011 | 0.80 | (0.51 – 1.24) | 0.31 | 0.60 | (0.41 – 0.88) | $9.6\times10^{-3}$ |
| Platelets | IVW-MRE | 668 | 0.98 | (0.91 – 1.05) | 0.53 | 0.90 | (0.80 – 1.01) | 0.071 | 1.03 | (0.94 – 1.12) | 0.54 |
| | Maximum likelihood | | 0.98 | (0.91 – 1.04) | 0.48 | 0.90 | (0.81 – 0.99) | 0.038 | 1.03 | (0.95 – 1.12) | 0.52 |
| | Weighted median | | 0.96 | (0.89 – 1.04) | 0.34 | 0.89 | (0.78 – 1.00) | 0.058 | 1.02 | (0.92 – 1.12) | 0.74 |
| | RAPS | | 0.97 | (0.90 – 1.06) | 0.53 | 0.90 | (0.79 – 1.02) | 0.097 | 1.02 | (0.93 – 1.12) | 0.66 |
| | PRESSO | | 0.97 | (0.91 – 1.04) | 0.45 | 0.90 | (0.80 – 1.00) | 0.049 | 1.02 | (0.94 – 1.12) | 0.58 |
| LMR | IVW-MRE | 450 | 1.05 | (0.80 – 1.37) | 0.73 | 1.11 | (0.77 – 1.60) | 0.58 | 1.02 | (0.74 – 1.39) | 0.91 |
| | Maximum likelihood | | 1.05 | (0.85 – 1.29) | 0.66 | 1.11 | (0.82 – 1.51) | 0.51 | 1.02 | (0.79 – 1.32) | 0.89 |
| | Weighted median | | 0.98 | (0.68 – 1.40) | 0.90 | 0.91 | (0.53 – 1.56) | 0.74 | 1.20 | (0.75 – 1.94) | 0.45 |
| | RAPS | | 0.94 | (0.75 – 1.19) | 0.61 | 0.87 | (0.63 – 1.21) | 0.41 | 1.01 | (0.75 – 1.36) | 0.96 |
| | PRESSO | | 0.93 | (0.75 – 1.15) | 0.49 | 0.95 | (0.69 – 1.29) | 0.73 | 0.92 | (0.69 – 1.21) | 0.54 |
| NLR | IVW-MRE | 249 | 1.17 | (0.83 – 1.66) | 0.37 | 1.06 | (0.64 – 1.76) | 0.82 | 1.29 | (0.85 – 1.95) | 0.24 |
| | Maximum likelihood | | 1.18 | (0.86 – 1.61) | 0.32 | 1.06 | (0.66 – 1.69) | 0.81 | 1.29 | (0.87 – 1.92) | 0.20 |
| | Weighted median | | 1.21 | (0.72 – 2.02) | 0.47 | 0.98 | (0.46 – 2.07) | 0.96 | 1.62 | (0.85 – 3.12) | 0.14 |
| | RAPS | | 1.17 | (0.81 – 1.70) | 0.39 | 1.11 | (0.64 – 1.90) | 0.72 | 1.26 | (0.80 – 1.96) | 0.32 |
| | PRESSO | | 1.17 | (0.83 – 1.66) | 0.37 | 1.06 | (0.64 – 1.77) | 0.82 | 1.29 | (0.85 – 1.95) | 0.24 |
| PLR | IVW-MRE | 470 | 1.38 | (1.09 – 1.75) | $7.2\times10^{-3}$ | 1.15 | (0.82 – 1.61) | 0.43 | 1.53 | (1.15 – 2.03) | $3.7\times10^{-3}$ |
| | Maximum likelihood | | 1.39 | (1.13 – 1.71) | $1.7\times10^{-3}$ | 1.15 | (0.85 – 1.55) | 0.37 | 1.54 | (1.19 – 1.98) | $9.4\times10^{-4}$ |
| | Weighted median | | 1.25 | (0.88 – 1.78) | 0.21 | 1.17 | (0.71 – 1.94) | 0.53 | 1.47 | (0.97 – 2.24) | 0.072 |
| | RAPS | | 1.38 | (1.08 – 1.76) | 0.011 | 1.19 | (0.84 – 1.70) | 0.33 | 1.48 | (1.10 – 2.00) | $9.6\times10^{-3}$ |
| | PRESSO | | 1.40 | (1.11 – 1.77) | $4.9\times10^{-3}$ | 1.14 | (0.82 – 1.61) | 0.43 | 1.55 | (1.17 – 2.06) | $2.4\times10^{-3}$ |

Abbreviations:

| | |
|---|---|
| IVW-MRE | Inverse variance weighted multiplicative random effect |
| RAPS | Robust adjusted profile score |
| PRESSO | Pleiotropy Residual Sum and Outlier |
| LMR | Lymphocyte to monocyte ratio |
| NLR | Neutrophil to lymphocyte ratio |
| PLR | Platelet to lymphocyte ratio |

**Supplementary Table S4: Mendelian randomization results for IDH wildtype ($IDH_{wt}$) glioma.** Odds ratios (OR) and 95% confidence intervals (CI) for the effect of increasing blood cell counts or cell type ratios on disease risk. MR Egger results are only presented for phenotypes with evidence of directional horizontal pleiotropy (MR Egger intercept $\beta_0 \neq 0$ (p<0.05).

| Blood Trait | MR Estimator | $N_{SNP}$ | $IDH_{wt}$ | | |
|---|---|---|---|---|---|
| | | | OR | (95% CI) | P |
| Basophils | IVW-MRE | 146 | 0.87 | (0.56 – 1.36) | 0.54 |
| | Maximum likelihood | | 0.87 | (0.55 – 1.37) | 0.54 |
| | Weighted median | | 0.81 | (0.40 – 1.62) | 0.55 |
| | RAPS | | 0.86 | (0.54 – 1.38) | 0.53 |
| | PRESSO | | 0.87 | (0.56 – 1.36) | 0.54 |
| Eosinophils | IVW-MRE | 370 | 0.98 | (0.80 – 1.19) | 0.81 |
| | Maximum likelihood | | 0.98 | (0.80 – 1.19) | 0.81 |
| | Weighted median | | 1.13 | (0.83 – 1.53) | 0.45 |
| | RAPS | | 1.02 | (0.82 – 1.25) | 0.88 |
| | PRESSO | | 0.98 | (0.80 – 1.19) | 0.81 |
| Lymphocytes | IVW-MRE | 414 | 1.05 | (0.89 – 1.23) | 0.59 |
| | Maximum likelihood | | 1.05 | (0.89 – 1.23) | 0.58 |
| | Weighted median | | 1.19 | (0.85 – 1.67) | 0.31 |
| | RAPS | | 1.05 | (0.88 – 1.25) | 0.57 |
| | PRESSO | | 1.05 | (0.89 – 1.23) | 0.59 |
| Monocytes | IVW-MRE | 479 | 1.04 | (1.00 – 1.07) | 0.058 |
| | Maximum likelihood | | 1.04 | (1.00 – 1.07) | 0.052 |
| | Weighted median | | 1.04 | (1.00 – 1.08) | 0.036 |
| | RAPS | | 1.04 | (1.00 – 1.08) | 0.056 |
| | PRESSO | | 1.04 | (1.00 – 1.07) | 0.058 |
| Neutrophils | IVW-MRE | 296 | 1.03 | (0.78 – 1.38) | 0.82 |
| | Maximum likelihood | | 1.03 | (0.82 – 1.31) | 0.78 |
| | Weighted median | | 0.87 | (0.60 – 1.26) | 0.45 |
| | RAPS | | 1.00 | (0.76 – 1.30) | 0.98 |
| | PRESSO | | 0.94 | (0.74 – 1.21) | 0.65 |
| Platelets | IVW-MRE | 668 | 1.03 | (0.97 – 1.10) | 0.37 |
| | Maximum likelihood | | 1.03 | (0.97 – 1.09) | 0.33 |
| | Weighted median | | 1.00 | (0.94 – 1.07) | 0.93 |
| | RAPS | | 1.03 | (0.96 – 1.09) | 0.42 |
| | PRESSO | | 1.02 | (0.96 – 1.08) | 0.49 |
| LMR | IVW-MRE | 450 | 1.08 | (0.89 – 1.30) | 0.44 |
| | Maximum likelihood | | 1.08 | (0.90 – 1.29) | 0.41 |
| | Weighted median | | 1.01 | (0.74 – 1.38) | 0.96 |
| | RAPS | | 1.03 | (0.85 – 1.26) | 0.76 |

| | | | | | |
|---|---|---|---|---|---|
| | PRESSO | | 1.08 | (0.90 – 1.29) | 0.44 |
| NLR | IVW-MRE | 249 | 0.96 | (0.71 – 1.30) | 0.81 |
| | Maximum likelihood | | 0.96 | (0.74 – 1.26) | 0.79 |
| | Weighted median | | 0.97 | (0.64 – 1.48) | 0.90 |
| | RAPS | | 0.93 | (0.69 – 1.25) | 0.62 |
| | MR Egger | | 0.42 | (0.18 – 0.93) | 0.034 |
| | PRESSO | | 0.89 | (0.68 – 1.18) | 0.43 |
| PLR | IVW-MRE | 470 | 1.23 | (1.01 – 1.49) | 0.036 |
| | Maximum likelihood | | 1.23 | (1.04 – 1.47) | 0.017 |
| | Weighted median | | 1.07 | (0.80 – 1.43) | 0.64 |
| | RAPS | | 1.21 | (1.00 – 1.47) | 0.051 |
| | MR Egger | | 0.83 | (0.53 – 1.30) | 0.42 |
| | PRESSO | | 1.19 | (1.00 – 1.42) | 0.055 |

Abbreviations:

IVW-MRE Inverse variance weighted multiplicative random effect

RAPS Robust adjusted profile score

PRESSO Pleiotropy Residual Sum and Outlier

LMR Lymphocyte to monocyte ratio

NLR Neutrophil to lymphocyte ratio

PLR Platelet to lymphocyte ratio

**Supplementary Table S5: Mendelian randomization results following manual removal of invalid genetic instruments.** Odds ratios (OR) and 95% for glioma for selected traits were estimated after filtering instruments that contributed to significant heterogeneity.

| Blood Cell Trait | Outcome | N / $N_{SNP}$ | IVW-MRE | | | Diagnostics | |
|---|---|---|---|---|---|---|---|
| | | | OR | (95% CI) | P | $P_{Q\text{-value}}$ | $P_{Egger}$ |
| PLR | Glioma overall | 442 / 470 | 1.27 | (1.12 – 1.44) | $2.2\times10^{-4}$ | 0.98 | 0.18 |
| PLR | $IDH_{mut}$ | 425 / 470 | 1.45 | (1.18 – 1.77) | $3.1\times10^{-4}$ | 0.99 | 0.40 |
| PLR | 1p/19q non-codeleted ($IDH_{mut\text{-}noncodel}$) | 434 / 470 | 1.53 | (1.19 – 1.96) | $8.1\times10^{-4}$ | 0.99 | 0.11 |
| Lymphocytes | $IDH_{mut}$ | 388 / 414 | 0.68 | (0.55 – 0.83) | $1.8\times10^{-4}$ | 0.99 | 0.52 |
| Lymphocytes | 1p/19q non-codeleted ($IDH_{mut\text{-}noncodel}$) | 391 / 414 | 0.69 | (0.55 – 0.85) | $4.5\times10^{-4}$ | 0.99 | 0.20 |
| Neutrophils | $IDH_{mut}$ | 271 / 296 | 0.65 | (0.50 – 0.85) | $1.4\times10^{-3}$ | 0.97 | 0.16 |
| Neutrophils | 1p/19q non-codeleted ($IDH_{mut\text{-}noncodel}$) | 278 / 296 | 0.61 | (0.43 – 0.86) | $5.2\times10^{-3}$ | 0.73 | 0.51 |

Abbreviations:

IVW-MRE  Inverse variance weighted multiplicative random effect

PLR  Platelet to lymphocyte ratio

**Supplementary Table S6: Multivariable (MV) Mendelian randomization results.** Odds ratios (OR) and 95% for glioma, estimated using MVMR for traits that were individually associated with specific glioma subtypes or were used to derive associated ratio phenotypes.

| Outcome | Traits Included | MV Method | Association Estimates | | |
|---|---|---|---|---|---|
| | | | OR | (95% CI) | P |
| $IDH_{mut}$ | Lymphocytes | IVW | 0.78 | (0.58 – 1.06) | 0.11 |
| | Neutrophils | IVW | 1.08 | (0.91 – 1.28) | 0.37 |
| | Platelets | IVW | 1.09 | (0.91 – 1.30) | 0.36 |
| | PLR | LASSO | 1.11 | | |
| | Lymphocytes | LASSO | - | | |
| | Neutrophils | LASSO | 0.86 | | |
| | Platelets | LASSO | - | | |
| $IDH_{mut-noncodel}$ | Lymphocytes | IVW | 0.70 | (0.48 – 1.00) | 0.051 |
| | Neutrophils | IVW | 1.12 | (0.91 – 1.40) | 0.27 |
| | Platelets | IVW | 1.10 | (0.89 – 1.37) | 0.36 |
| | PLR | LASSO | 1.16 | | |
| | Lymphocytes | LASSO | 0.89 | | |
| | Neutrophils | LASSO | 0.90 | | |
| | Platelets | LASSO | - | | |

**Supplementary Table S7: Candidate variants for colocalization.** Genome-wide significant glioma risk variants identified among genetic instruments for blood cell traits. For each variant associations with all blood cell phenotypes and/or leukocyte telomere length that reached genome-wide significance are shown.

| Region | SNP[1] | Outcome | OR | P-value | P-values for associated traits | |
|---|---|---|---|---|---|---|
| 2q37.3 | rs34290285 (G/A) | $IDH_{mut}$ 1p/19q non-codel | 0.60 | $5.2\times10^{-10}$ | Eosinophils: | $3.9\times10^{-37}$ |
| | | $IDH_{mut}$ | 0.66 | $1.6\times10^{-9}$ | | |
| 5p15.33 | rs7705526 (C/A) | $IDH_{wt}$ | 0.60 | $6.7\times10^{-25}$ | Neutrophils:<br>NLR:<br>PLR:<br>Platelets:<br>Telomere length: | $1.3\times10^{-12}$<br>$1.4\times10^{-9}$<br>$2.7\times10^{-12}$<br>$9.9\times10^{-33}$<br>$2.4\times10^{-282}$ |
| | | $IDH_{wt}$ | 0.60 | $6.7\times10^{-25}$ | | |
| | | Glioma overall | 0.68 | $2.2\times10^{-23}$ | | |
| 5p15.33 | rs2853677 (G/A) | Glioma overall | 1.25 | $1.2\times10^{-9}$ | Neutrophils:<br>NLR:<br>PLR:<br>Platelets:<br>Telomere length: | $2.2\times10^{-11}$<br>$5.7\times10^{-14}$<br>$3.2\times10^{-12}$<br>$3.4\times10^{-21}$<br>$3.1\times10^{-127}$ |
| 8q24 | rs72716319 (A/G) | $IDH_{mut}$ | 0.27 | $2.5\times10^{-41}$ | LMR:<br>Monocytes: | $5.3\times10^{-21}$<br>$4.0\times10^{-21}$ |
| | | $IDH_{mut}$ 1p/19q non-codel | 0.27 | $5.2\times10^{-29}$ | | |
| | | Glioma overall | 0.47 | $6.2\times10^{-24}$ | | |
| | | $IDH_{mut}$ 1p/19q co-del | 0.26 | $1.5\times10^{-23}$ | | |
| 8q24 | rs55705857 (A/G) | $IDH_{mut}$ | 0.24 | $3.0\times10^{-55}$ | LMR:<br>Monocytes: | $1.0\times10^{-22}$<br>$1.6\times10^{-19}$ |
| | | $IDH_{mut}$ 1p/19q co-del | 0.20 | $8.9\times10^{-37}$ | | |
| | | $IDH_{mut}$ 1p/19q non-codel | 0.25 | $2.7\times10^{-34}$ | | |
| | | Glioma overall | 0.44 | $3.1\times10^{-32}$ | | |
| 8q24 | rs16904140 (G/A) | $IDH_{mut}$ | 0.57 | $1.3\times10^{-19}$ | LMR:<br>Monocytes:<br>Platelets: | $6.1\times10^{-21}$<br>$2.2\times10^{-17}$<br>$1.1\times10^{-8}$ |
| | | $IDH_{mut}$ 1p/19q co-del | 0.48 | $2.0\times10^{-17}$ | | |
| | | Glioma overall | 0.76 | $1.5\times10^{-10}$ | | |
| | | $IDH_{mut}$ 1p/19q non-codel | 0.64 | $4.8\times10^{-9}$ | | |
| 20q13.33 | rs4809319 (A/G) | $IDH_{wt}$ | 0.62 | $1.3\times10^{-14}$ | Platelets:<br>Telomere length: | $1.1\times10^{-9}$<br>$1.1\times10^{-19}$ |
| | | Glioma overall | 0.71 | $1.4\times10^{-13}$ | | |

[1] Alleles are listed as effect allele/other allele, where the odds ratio corresponds to the effect allele

**Supplementary Table S8: Colocalization results.** Regions with evidence of colocalization, denoted by posterior probability (PP)>0.90 between signals for glioma susceptibility and putative risk factors.

| Locus | Candidate | Colocalized Traits | Dropped Traits | PP[1] | PP$_{regional}$[2] | Fine-mapped[3] | PP$_{SNP}$ |
|---|---|---|---|---|---|---|---|
| 2q37.3 | rs34290285 (*D2HGDH*) | IDH$_{mut}$ 1p/19q non-codel, Eosinophils | - | 0.939 | 1 | rs34290285 | 0.944 |
| 5p15.33 | rs7705526 (*TERT*) | IDH$_{wt}$, telomere length, platelets, neutrophils, PLR | - | 0.994 | 1 | rs7705526 | 1 |
| 5p15.33 | rs2853677 (*TERT*) | Glioma, telomere length, platelets | - | 0.978 | 1 | rs7705526 | 1 |
| 20q13.33 | rs4809319 (*STMN3*) | None | Telomere length | - | 0.665 | - | - |
| | | IDH$_{wt}$, platelets | - | 0.952 | 1 | rs6011018 (*RTEL1*) | 0.178 |

[1] Posterior probability that all traits are colocalized

[2] Posterior probability that one or more SNPs in the region have shared associations across the traits

[3] Candidate variant explaining the shared association

[4] Proportion of the posterior probability explained by the fine-mapped variant

Abbreviations:

PP Posterior probability

**Supplementary Table S9: Survival analysis of blood cell trait polygenic scores (PGS).** Hazard ratios (HR) and 95% confidence intervals per 1 standard deviation increase in the standardized PGS were estimated using Cox proportional hazards models fit separately in each patient population. Associations of each PGS with all-cause mortality were estimated with adjustment for age, sex, and top 10 genetic ancestry principal components.

| Blood Cell PGS | Cases (Events) | UCSF AGS and Mayo Clinic | | | Cases (Events) | UCSF AGS | | | Cases (Events) | TCGA | | |
|---|---|---|---|---|---|---|---|---|---|---|---|---|
| | | HR | (95% CI) | P | | HR | (95% CI) | P | | HR | (95% CI) | P |
| Basophils | Glioma 1973 (1218) | 1.02 | (0.97 – 1.08) | 0.43 | Glioma 659 (592) | 1.01 | (0.93 – 1.09) | 0.83 | Glioma 786 (310) | 1.14 | (1.00 – 1.14) | 0.042 |
| Eosinophils | | 0.99 | (0.93 – 1.05) | 0.73 | | 1.04 | (0.96 – 1.13) | 0.30 | | 1.03 | (0.92 – 1.16) | 0.64 |
| Lymphocytes | | 1.02 | (0.96 – 1.08) | 0.48 | | 1.00 | (0.93 – 1.09) | 0.94 | | 1.14 | (1.01 – 1.29) | 0.030 |
| Monocytes | | 1.00 | (0.95 – 1.06) | 0.89 | | 1.01 | (0.93 – 1.10) | 0.83 | | 0.93 | (0.83 – 1.03) | 0.17 |
| Neutrophils | | 1.05 | (0.99 – 1.11) | 0.12 | | 0.97 | (0.90 – 1.05) | 0.43 | | 1.07 | (0.95 – 1.21) | 0.26 |
| Platelets | | 1.01 | (0.96 – 1.07) | 0.65 | | 1.05 | (0.97 – 1.14) | 0.23 | | 0.98 | (0.88 – 1.09) | 0.72 |
| LMR | | 0.96 | (0.91 – 1.02) | 0.21 | | 1.00 | (0.92 – 1.09) | 0.98 | | 1.10 | (0.99 – 1.23) | 0.090 |
| NLR | | 1.00 | (0.95 – 1.06) | 0.94 | | 0.96 | (0.88 – 1.04) | 0.29 | | 0.87 | (0.78 – 0.97) | 0.013 |
| PLR | | 0.97 | (0.91 – 1.02) | 0.24 | | 1.00 | (0.92 – 1.08) | 0.97 | | 0.95 | (0.85 – 1.06) | 0.36 |
| Basophils | $IDH_{mut}$ 588 (201) | 1.11 | (0.96 – 1.28) | 0.18 | $IDH_{mut}$ 111 (74) | 1.00 | (0.78 – 1.29) | 0.99 | $IDH_{mut}$ 375 (50) | 1.70 | (1.22 – 2.37) | 1.8E-03 |
| Eosinophils | | 1.17 | (1.00 – 1.37) | 0.055 | | 0.89 | (0.70 – 1.14) | 0.36 | | 1.18 | (0.86 – 1.61) | 0.31 |
| Lymphocytes | | 1.09 | (0.94 – 1.27) | 0.26 | | 0.99 | (0.78 – 1.25) | 0.91 | | 1.42 | (1.03 – 1.95) | 0.030 |
| Monocytes | | 0.98 | (0.85 – 1.13) | 0.79 | | 0.79 | (0.59 – 1.06) | 0.11 | | 0.86 | (0.63 – 1.18) | 0.35 |
| Neutrophils | | 1.12 | (0.96 – 1.31) | 0.15 | | 0.95 | (0.76 – 1.19) | 0.66 | | 1.35 | (1.00 – 1.82) | 0.051 |
| Platelets | | 1.04 | (0.90 – 1.20) | 0.58 | | 0.96 | (0.76 – 1.21) | 0.72 | | 1.09 | (0.83 – 1.45) | 0.53 |
| LMR | | 0.93 | (0.80 – 1.07) | 0.30 | | 1.06 | (0.83 – 1.36) | 0.64 | | 1.32 | (0.96 – 1.81) | 0.083 |
| NLR | | 0.91 | (0.80 – 1.05) | 0.20 | | 0.95 | (0.76 – 1.19) | 0.67 | | 0.76 | (0.56 – 1.03) | 0.073 |
| PLR | | 0.93 | (0.81 – 1.08) | 0.34 | | 0.88 | (0.70 – 1.10) | 0.25 | | 0.84 | (0.60 – 1.16) | 0.29 |
| Basophils | | 1.33 | (1.00 – 1.77) | 0.047 | | - | - | - | | 2.59 | (1.04 – 6.41) | 0.040 |
| Eosinophils | | 1.56 | (1.15 – 2.13) | $4.4 \times 10^{-3}$ | | - | - | - | | 1.94 | (0.60 – 6.32) | 0.27 |

| | | | | | | | | | | | | |
|---|---|---|---|---|---|---|---|---|---|---|---|---|
| Lymphocytes | $IDH_{mut}$ 1p/19q co-del 244 (64) | 1.68 | (1.24 – 2.27) | $7.4\times10^{-4}$ | $IDH_{mut}$ 1p/19q co-del 9 (1) | - | - | - | $IDH_{mut}$ 1p/19q co-del 143 (13) | 1.38 | (0.50 – 3.81) | 0.53 |
| Monocytes | | 1.22 | (0.94 – 1.59) | 0.13 | | - | - | - | | 1.71 | (0.65 – 4.48) | 0.28 |
| Neutrophils | | 1.37 | (1.03 – 1.83) | 0.033 | | - | - | - | | 4.82 | (1.63 – 14.29) | 4.5E-03 |
| platelet | | 0.80 | (0.61 – 1.05) | 0.11 | | - | - | - | | 1.50 | (0.59 – 3.80) | 0.39 |
| LMR | | 0.99 | (0.77 – 1.29) | 0.97 | | - | - | - | | 1.42 | (0.58 – 3.46) | 0.44 |
| NLR | | 0.83 | (0.64 – 1.07) | 0.15 | | - | - | - | | 1.78 | (0.73 – 4.35) | 0.20 |
| PLR | | 0.83 | (0.63 – 1.08) | 0.17 | | - | - | - | | 0.87 | (0.40 – 1.91) | 0.73 |
| Basophils | $IDH_{mut}$ 1p/19q non-codel 291 (117) | 1.13 | (0.93 – 1.39) | 0.22 | $IDH_{mut}$ 1p/19q non-codel 94 (69) | 1.09 | (0.84 – 1.41) | 0.51 | $IDH_{mut}$ 1p/19q non-codel 230 (36) | 1.63 | (1.08 – 2.45) | 0.019 |
| Eosinophils | | 1.08 | (0.88 – 1.32) | 0.46 | | 0.97 | (0.75 – 1.25) | 0.79 | | 1.15 | (0.81 – 1.64) | 0.43 |
| Lymphocytes | | 1.02 | (0.84 – 1.24) | 0.86 | | 1.11 | (0.87 – 1.41) | 0.41 | | 1.33 | (0.85 – 2.07) | 0.21 |
| Monocytes | | 1.04 | (0.85 – 1.28) | 0.67 | | 0.87 | (0.64 – 1.20) | 0.41 | | 0.64 | (0.42 – 0.99) | 0.045 |
| Neutrophils | | 1.03 | (0.85 – 1.26) | 0.75 | | 1.03 | (0.83 – 1.28) | 0.78 | | 1.14 | (0.77 – 1.68) | 0.51 |
| Platelets | | 1.04 | (0.86 – 1.27) | 0.67 | | 0.90 | (0.70 – 1.15) | 0.41 | | 0.99 | (0.70 – 1.39) | 0.94 |
| LMR | | 0.95 | (0.79 – 1.14) | 0.57 | | 1.11 | (0.85 – 1.43) | 0.44 | | 1.35 | (0.92 – 1.96) | 0.12 |
| NLR | | 0.88 | (0.73 – 1.07) | 0.19 | | 0.89 | (0.71 – 1.11) | 0.29 | | 0.70 | (0.43 – 1.13) | 0.15 |
| PLR | | 0.96 | (0.80 – 1.15) | 0.66 | | 0.83 | (0.65 – 1.06) | 0.13 | | 0.83 | (0.53 – 1.30) | 0.42 |
| Basophils | $IDH_{wt}$ 699 (594) | 1.00 | (0.93 – 1.09) | 0.91 | $IDH_{wt}$ 416 (402) | 1.04 | (0.94 – 1.14) | 0.44 | $IDH_{wt}$ 364 (228) | 1.11 | (0.95 – 1.29) | 0.18 |
| Eosinophils | | 0.95 | (0.87 – 1.04) | 0.25 | | 1.07 | (0.97 – 1.18) | 0.18 | | 1.09 | (0.95 – 1.25) | 0.23 |
| Lymphocytes | | 1.04 | (0.96 – 1.13) | 0.32 | | 1.01 | (0.91 – 1.11) | 0.91 | | 1.09 | (0.94 – 1.26) | 0.23 |
| Monocytes | | 0.98 | (0.91 – 1.06) | 0.64 | | 1.03 | (0.93 – 1.13) | 0.57 | | 0.89 | (0.78 – 1.01) | 0.073 |
| Neutrophils | | 1.02 | (0.94 – 1.10) | 0.69 | | 0.99 | (0.90 – 1.08) | 0.81 | | 0.99 | (0.86 – 1.15) | 0.91 |
| Platelets | | 1.03 | (0.95 – 1.11) | 0.53 | | 1.10 | (1.00 – 1.22) | 0.061 | | 0.90 | (0.79 – 1.02) | 0.11 |
| LMR | | 1.00 | (0.92 – 1.09) | 0.96 | | 0.95 | (0.86 – 1.05) | 0.33 | | 1.09 | (0.96 – 1.24) | 0.16 |
| NLR | | 1.01 | (0.93 – 1.10) | 0.85 | | 0.99 | (0.89 – 1.10) | 0.86 | | 0.92 | (0.81 – 1.04) | 0.16 |
| PLR | | 0.95 | (0.87 – 1.03) | 0.20 | | 1.04 | (0.94 – 1.15) | 0.44 | | 0.92 | (0.80 – 1.05) | 0.19 |

**Supplementary Table S10: Meta-analysis of blood cell trait polygenic scores (PGS) associations.** Study specific results in Supplementary Table S9 were combined in a fixed-effects meta-analysis to estimate the overall effect of each blood cell trait PGS on survival. Hazard ratios (HR) correspond to a 1 standard deviation increase in the standardized PGS.

| Blood Cell PGS | Cases (Events) | Meta-Analysis Estimates | | | | Heterogeneity[2] | |
|---|---|---|---|---|---|---|---|
| | | HR | (95% CI) | P | FDR[1] | Q P-value | $I^2$ |
| Basophils | Glioma: 3418 (2120) | 1.03 | (0.99 – 1.08) | 0.15 | 0.47 | 0.25 | 0.27 |
| Eosinophils | | 1.01 | (0.97 – 1.06) | 0.62 | 0.79 | 0.56 | 0 |
| Lymphocytes | | 1.03 | (0.99 – 1.08) | 0.17 | 0.47 | 0.19 | 0.41 |
| Monocytes | | 0.99 | (0.95 – 1.04) | 0.75 | 0.80 | 0.39 | 0 |
| Neutrophils | | 1.03 | (0.98 – 1.07) | 0.26 | 0.47 | 0.21 | 0.36 |
| Platelets | | 1.02 | (0.98 – 1.06) | 0.41 | 0.61 | 0.57 | 0 |
| LMR | | 0.99 | (0.95 – 1.04) | 0.80 | 0.80 | 0.11 | 0.54 |
| NLR | | 0.97 | (0.93 – 1.01) | 0.14 | 0.47 | 0.079 | 0.61 |
| PLR | | 0.97 | (0.93 – 1.02) | 0.21 | 0.47 | 0.72 | 0 |
| Basophils | $IDH_{mut}$ 1074 (325) | 1.14 | (1.02 – 1.29) | 0.027 | 0.19 | 0.035 | 0.70 |
| Eosinophils | | 1.09 | (0.97 – 1.24) | 0.16 | 0.24 | 0.17 | 0.44 |
| Lymphocytes | | 1.10 | (0.98 – 1.24) | 0.10 | 0.19 | 0.19 | 0.40 |
| Monocytes | | 0.93 | (0.82 – 1.05) | 0.22 | 0.28 | 0.37 | 0 |
| Neutrophils | | 1.10 | (0.98 – 1.24) | 0.11 | 0.19 | 0.18 | 0.42 |
| Platelets | | 1.03 | (0.92 – 1.15) | 0.60 | 0.68 | 0.75 | 0 |
| LMR | | 1.00 | (0.89 – 1.12) | 0.99 | 1.00 | 0.11 | 0.54 |
| NLR | | 0.90 | (0.81 – 1.01) | 0.063 | 0.19 | 0.46 | 0 |
| PLR | | 0.91 | (0.81 – 1.02) | 0.089 | 0.19 | 0.79 | 0 |
| Basophils | $IDH_{mut\text{-}codel}$ 396 (78) | 1.42 | (1.08 – 1.86) | 0.012 | 0.03 | 0.17 | 0.46 |
| Eosinophils | | 1.59 | (1.18 – 2.14) | $2.4\times10^{-3}$ | 0.011 | 0.73 | 0 |
| Lymphocytes | | 1.65 | (1.24 – 2.20) | $6.4\times10^{-4}$ | $5.8\times10^{-3}$ | 0.72 | 0 |
| Monocytes | | 1.25 | (0.97 – 1.61) | 0.079 | 0.14 | 0.51 | 0 |
| Neutrophils | | 1.49 | (1.13 – 1.97) | $5.2\times10^{-3}$ | 0.016 | 0.029 | 0.79 |
| platelet | | 0.84 | (0.65 – 1.09) | 0.20 | 0.25 | 0.20 | 0.38 |
| LMR | | 1.02 | (0.80 – 1.32) | 0.85 | 0.85 | 0.45 | 0 |
| NLR | | 0.88 | (0.69 – 1.13) | 0.31 | 0.34 | 0.11 | 0.62 |
| PLR | | 0.83 | (0.64 – 1.07) | 0.16 | 0.23 | 0.90 | 0 |
| Basophils | | 1.17 | (1.01 – 1.36) | 0.033 | 0.30 | 0.23 | 0.32 |
| Eosinophils | | 1.05 | (0.91 – 1.22) | 0.48 | 0.68 | 0.68 | 0 |

| | | | | | | | |
|---|---|---|---|---|---|---|---|
| Lymphocytes | IDHmut-noncodel 615 (222) | 1.08 | (0.93 – 1.25) | 0.31 | 0.68 | 0.55 | 0 |
| Monocytes | | 0.93 | (0.80 – 1.10) | 0.41 | 0.68 | 0.12 | 0.52 |
| Neutrophils | | 1.04 | (0.91 – 1.20) | 0.53 | 0.68 | 0.90 | 0 |
| Platelets | | 0.99 | (0.86 – 1.14) | 0.84 | 0.95 | 0.66 | 0 |
| LMR | | 1.00 | (0.87 – 1.15) | 0.98 | 0.98 | 0.10 | 0.57 |
| NLR | | 0.90 | (0.79 – 1.03) | 0.14 | 0.43 | 0.49 | 0 |
| PLR | | 0.90 | (0.78 – 1.04) | 0.14 | 0.43 | 0.60 | 0 |
| Basophils | IDHwt 1479 (1224) | 1.03 | (0.97 – 1.09) | 0.30 | 0.71 | 0.52 | 0 |
| Eosinophils | | 1.02 | (0.96 – 1.08) | 0.59 | 0.76 | 0.12 | 0.53 |
| Lymphocytes | | 1.04 | (0.98 – 1.10) | 0.21 | 0.71 | 0.64 | 0 |
| Monocytes | | 0.98 | (0.93 – 1.03) | 0.44 | 0.71 | 0.21 | 0.36 |
| Neutrophils | | 1.00 | (0.95 – 1.06) | 0.93 | 0.93 | 0.89 | 0 |
| Platelets | | 1.02 | (0.97 – 1.08) | 0.44 | 0.71 | 0.052 | 0.66 |
| LMR | | 1.01 | (0.95 – 1.07) | 0.81 | 0.91 | 0.24 | 0.31 |
| NLR | | 0.98 | (0.92 – 1.04) | 0.47 | 0.71 | 0.48 | 0 |
| PLR | | 0.97 | (0.92 – 1.03) | 0.31 | 0.71 | 0.24 | 0.30 |

[1] False discovery rate (FDR) was calculated for each glioma subtype, FDR<0.05 was considered statistically significant

[2] Heterogeneity in study-specific associations was assessed using the standard Cochran's Q test ($p<0.05$ were considered statistically significant) and Higgins $I^2$ statistic, which describes the percentage of variation across studies that is due to heterogeneity rather than chance

**Supplementary Table S11: Mendelian randomization results for survival in *IDH*-mutated 1p19q co-deleted subtype.** Hazard ratios (HR) and 95% confidence intervals (CI) correspond to the effect of increasing blood cell counts or cell type ratios on all-cause mortality. Results are presented for analyses suing SNP effects on survival before and after index event bias correction. MR Egger estimates are presented if applicable (Egger intercept test p<0.05).

| Blood Trait | MR Estimator | $N_{SNP}$[1] | Unadjusted | | | Adjusted for Index Event Bias | | |
|---|---|---|---|---|---|---|---|---|
| | | | HR | (95% CI) | P | HR | (95% CI) | P |
| Basophils | IVW-MRE | 143 | 13.98 | (1.63 – 120.3) | 0.016 | 13.18 | (1.55 – 112.2) | 0.018 |
| | Maximum likelihood | | 14.00 | (2.24 – 87.8) | $4.8 \times 10^{-3}$ | 13.56 | (2.16 – 85.2) | $5.4 \times 10^{-3}$ |
| | Weighted median | | 46.24 | (2.10 – 1018.1) | 0.015 | 32.02 | (1.32 – 779.0) | 0.033 |
| | RAPS | | 16.48 | (1.72 – 158.1) | 0.015 | 15.05 | (1.57 – 144.5) | 0.019 |
| | PRESSO | | 13.98 | (1.63 – 120.3) | 0.017 | 13.18 | (1.55 – 112.2) | 0.020 |
| Eosinophils | IVW-MRE | 369 | 3.83 | (1.63 – 9.00) | $2.0 \times 10^{-3}$ | 3.87 | (1.65 – 9.09) | $1.9 \times 10^{-3}$ |
| | Maximum likelihood | | 3.82 | (1.74 – 8.39) | $8.5 \times 10^{-4}$ | 3.84 | (1.74 – 8.45) | $8.4 \times 10^{-4}$ |
| | Weighted median | | 5.15 | (1.35 – 19.68) | 0.017 | 5.27 | (1.35 – 20.62) | 0.017 |
| | RAPS | | 3.67 | (1.47 – 9.14) | $5.3 \times 10^{-3}$ | 3.68 | (1.47 – 9.20) | $5.4 \times 10^{-3}$ |
| | PRESSO | | 3.83 | (1.63 – 9.00) | $2.2 \times 10^{-3}$ | 3.87 | (1.65 – 9.09) | $2.1 \times 10^{-3}$ |
| Lymphocytes | IVW-MRE | 411 | 4.33 | (2.16 – 8.65) | $3.4 \times 10^{-5}$ | 4.36 | (2.18 – 8.73) | $3.2 \times 10^{-5}$ |
| | Maximum likelihood | | 4.40 | (2.21 – 8.77) | $2.6 \times 10^{-5}$ | 4.41 | (2.21 – 8.78) | $2.5 \times 10^{-5}$ |
| | Weighted median | | 1.27 | (0.36 – 4.43) | 0.71 | 1.32 | (0.36 – 4.81 | 0.68 |
| | RAPS | | 5.71 | (2.68 – 12.17) | $6.6 \times 10^{-6}$ | 5.56 | (2.61 – 11.85) | $9.0 \times 10^{-6}$ |
| | PRESSO | | 4.32 | (2.16 – 8.65) | $4.2 \times 10^{-5}$ | 4.36 | (2.18 – 8.73) | $3.9 \times 10^{-5}$ |
| Monocytes | IVW-MRE | 473 | 1.13 | (0.97 – 1.32) | 0.11 | 1.13 | (0.97 – 1.32) | 0.11 |
| | Maximum likelihood | | 1.13 | (0.98 – 1.30) | 0.087 | 1.13 | (0.98 – 1.30) | 0.089 |
| | Weighted median | | 1.12 | (0.96 – 1.29) | 0.15 | 1.12 | (0.97 – 1.28) | 0.13 |
| | RAPS | | 1.13 | (0.97 – 1.32) | 0.12 | 1.13 | (0.97 – 1.32) | 0.12 |
| | PRESSO | | 1.13 | (0.97 – 1.32) | 0.11 | 1.13 | (0.97 – 1.32) | 0.11 |
| Neutrophils | IVW-MRE | 293 | 4.71 | (1.72 – 12.91) | $2.6 \times 10^{-3}$ | 4.51 | (1.66 – 12.26) | $3.2 \times 10^{-3}$ |
| | Maximum likelihood | | 4.70 | (1.85 – 11.96) | $1.2 \times 10^{-3}$ | 4.50 | (1.77 – 11.49) | $1.6 \times 10^{-3}$ |
| | Weighted median | | 5.33 | (1.21 – 23.46) | 0.027 | 4.96 | (1.07 – 22.98) | 0.041 |
| | RAPS | | 4.05 | (1.37 – 12.02) | 0.012 | 3.87 | (1.32 – 11.38) | 0.014 |
| | MR Egger | | 28.45 | (2.29 – 353.4) | $9.7 \times 10^{-3}$ | 26.94 | (2.22 – 327.5) | 0.010 |
| | PRESSO | | 4.71 | (1.72 – 12.91) | $2.8 \times 10^{-3}$ | 4.51 | (1.66 – 12.26) | $3.5 \times 10^{-3}$ |
| Platelets | IVW-MRE | 659 | 0.84 | (0.67 – 1.05) | 0.13 | 0.83 | (0.66 – 1.04) | 0.10 |
| | Maximum likelihood | | 0.86 | (0.69 – 1.06) | 0.16 | 0.84 | (0.67 – 1.04) | 0.11 |
| | Weighted median | | 0.77 | (0.58 – 1.03) | 0.076 | 0.75 | (0.57 – 1.00) | 0.052 |
| | RAPS | | 0.83 | (0.65 – 1.07) | 0.16 | 0.82 | (0.64 – 1.06) | 0.13 |
| | PRESSO | | 0.84 | (0.67 – 1.05) | 0.13 | 0.83 | (0.66 – 1.04) | 0.10 |
| LMR | IVW-MRE | 446 | 1.10 | (0.53 – 2.30) | 0.80 | 1.13 | (0.54 – 2.36) | 0.75 |
| | Maximum likelihood | | 1.10 | (0.54 – 2.23) | 0.80 | 1.13 | (0.55 – 2.30) | 0.74 |

| | | | | | | | | |
|---|---|---|---|---|---|---|---|---|
| | Weighted median | | 0.88 | (0.26 – 2.98) | 0.83 | 0.90 | (0.26 – 3.04) | 0.86 |
| | RAPS | | 1.02 | (0.48 – 2.20) | 0.95 | 1.06 | (0.49 – 2.29) | 0.88 |
| | PRESSO | | 1.10 | (0.53 – 2.30) | 0.80 | 1.13 | (0.54 – 2.36) | 0.75 |
| NLR | IVW-MRE | 248 | 0.50 | (0.16 – 1.56) | 0.23 | 0.50 | (0.16 – 1.58) | 0.24 |
| | Maximum likelihood | | 0.50 | (0.17 – 1.44) | 0.20 | 0.50 | (0.17 – 1.46) | 0.20 |
| | Weighted median | | 0.91 | (0.17 – 4.83) | 0.91 | 0.71 | (0.13 – 3.90) | 0.70 |
| | RAPS | | 0.47 | (0.14 – 1.54) | 0.21 | 0.48 | (0.15 – 1.60) | 0.23 |
| | PRESSO | | 0.50 | (0.16 – 1.56) | 0.23 | 0.50 | (0.16 – 1.58) | 0.24 |
| PLR | IVW-MRE | 465 | 0.50 | (0.24 – 1.04) | 0.065 | 0.51 | (0.25 – 1.07) | 0.074 |
| | Maximum likelihood | | 0.51 | (0.25 – 1.02) | 0.056 | 0.52 | (0.26 – 1.04) | 0.063 |
| | Weighted median | | 0.53 | (0.17 – 1.65) | 0.27 | 0.67 | (0.22 – 2.06) | 0.49 |
| | RAPS | | 0.50 | (0.23 – 1.09) | 0.082 | 0.52 | (0.24 – 1.12) | 0.10 |
| | PRESSO | | 0.50 | (0.24 – 1.04) | 0.066 | 0.51 | (0.25 – 1.07) | 0.074 |

[1] Number of instruments using in the survival analysis is smaller due to the removal of variants with abs(log(HR))>2.5

Abbreviations:

| | |
|---|---|
| IVW-MRE | Inverse variance weighted multiplicative random effect |
| RAPS | Robust adjusted profile score |
| PRESSO | Pleiotropy Residual Sum and Outlier |
| LMR | Lymphocyte to monocyte ratio |
| NLR | Neutrophil to lymphocyte ratio |
| PLR | Platelet to lymphocyte ratio |

**Supplementary Table S12: Associations between blood cell trait polygenic scores (PGS) and tumor immune microenvironment (TIME) features in TCGA.** Associations for selected blood cell trait PGS with heritable TIME traits identified in TCGA by Sayaman et al. (2021). Association analyses were conducted stratified by IDH mutation status. Each continuous TME phenotype was analyzed using linear regression with adjustment for age, sex, and genetic ancestry principal components. Heterogeneity in PGS effects by IDH status were assessed using Cochran's Q test.

| Module | TME Trait | PGS | IDH mutant | | | | IDH wildtype | | | | $P_{het}$ |
|---|---|---|---|---|---|---|---|---|---|---|---|
| | | | Beta | SE | P | FDR | Beta | SE | P | FDR | |
| Expression Signature (ES) | Bcell mg IGJ | Basophils | -0.008 | 0.034 | 0.809 | 0.839 | 0.038 | 0.067 | 0.572 | 0.854 | 0.539 |
| | | Eosinophils | -0.005 | 0.036 | 0.885 | 0.991 | 0.008 | 0.061 | 0.896 | 0.939 | 0.852 |
| | | Lymphocytes | 0.032 | 0.036 | 0.366 | 0.528 | -0.024 | 0.064 | 0.701 | 0.818 | 0.437 |
| | | Neutrophils | 0.062 | 0.035 | 0.077 | 0.197 | -0.009 | 0.072 | 0.901 | 0.970 | 0.372 |
| | | Platelets | -0.036 | 0.032 | 0.252 | 0.655 | -0.024 | 0.059 | 0.688 | 0.917 | 0.850 |
| | | PLR | -0.046 | 0.033 | 0.166 | 0.307 | 0.034 | 0.063 | 0.587 | 0.748 | 0.258 |
| | CD8 CD68 ratio | Basophils | -0.083 | 0.069 | 0.232 | 0.719 | 0.086 | 0.077 | 0.262 | 0.854 | 0.101 |
| | | Eosinophils | 0.080 | 0.073 | 0.274 | 0.991 | 0.006 | 0.069 | 0.936 | 0.939 | 0.460 |
| | | Lymphocytes | -0.052 | 0.072 | 0.472 | 0.574 | 0.010 | 0.073 | 0.894 | 0.900 | 0.547 |
| | | Neutrophils | -0.0002 | 0.072 | 0.997 | 0.997 | 0.022 | 0.082 | 0.788 | 0.964 | 0.838 |
| | | Platelets | -0.028 | 0.064 | 0.657 | 0.836 | -0.118 | 0.067 | 0.077 | 0.729 | 0.330 |
| | | PLR | 0.108 | 0.067 | 0.112 | 0.223 | -0.055 | 0.072 | 0.442 | 0.652 | 0.098 |
| | LYMPHS PCA 16704732 | Basophils | 0.027 | 0.020 | 0.179 | 0.719 | -0.026 | 0.040 | 0.514 | 0.854 | 0.235 |
| | | Eosinophils | 0.018 | 0.021 | 0.396 | 0.991 | -0.037 | 0.036 | 0.307 | 0.683 | 0.189 |
| | | Lymphocytes | 0.019 | 0.021 | 0.377 | 0.528 | -0.066 | 0.037 | 0.074 | 0.209 | 0.046 |
| | | Neutrophils | 0.026 | 0.021 | 0.221 | 0.428 | -0.060 | 0.042 | 0.155 | 0.964 | 0.068 |
| | | Platelets | 0.023 | 0.019 | 0.217 | 0.655 | 0.053 | 0.034 | 0.123 | 0.729 | 0.449 |
| | | PLR | -0.019 | 0.020 | 0.349 | 0.444 | 0.079 | 0.036 | 0.032 | 0.299 | 0.019 |
| | PD1 | Basophils | 0.041 | 0.053 | 0.443 | 0.719 | -0.057 | 0.072 | 0.427 | 0.854 | 0.273 |
| | | Eosinophils | 0.034 | 0.056 | 0.542 | 0.991 | -0.044 | 0.065 | 0.495 | 0.769 | 0.359 |
| | | Lymphocytes | 0.054 | 0.056 | 0.338 | 0.525 | -0.066 | 0.067 | 0.327 | 0.532 | 0.170 |
| | | Neutrophils | 0.129 | 0.055 | 0.019 | 0.090 | -0.054 | 0.075 | 0.476 | 0.964 | 0.049 |
| | | Platelets | -0.031 | 0.050 | 0.534 | 0.732 | -0.032 | 0.062 | 0.599 | 0.883 | 0.984 |

| | | | | | | | | | | | |
|---|---|---|---|---|---|---|---|---|---|---|---|
| | | PLR | -0.069 | 0.052 | 0.186 | 0.307 | 0.063 | 0.066 | 0.344 | 0.567 | 0.117 |
| IFN Response | Activated Dendritic Cells (ES) | Basophils | -0.004 | 0.034 | 0.903 | 0.903 | -0.019 | 0.055 | 0.723 | 0.854 | 0.813 |
| | | Eosinophils | -0.008 | 0.036 | 0.820 | 0.991 | -0.049 | 0.049 | 0.319 | 0.683 | 0.503 |
| | | Lymphocytes | 0.057 | 0.036 | 0.115 | 0.354 | -0.112 | 0.050 | 0.027 | 0.192 | 6.4E-03 |
| | | Neutrophils | 0.043 | 0.036 | 0.229 | 0.428 | -0.030 | 0.057 | 0.605 | 0.964 | 0.282 |
| | | Platelets | -0.056 | 0.032 | 0.082 | 0.655 | 0.004 | 0.047 | 0.932 | 0.988 | 0.294 |
| | | PLR | -0.091 | 0.033 | 0.007 | 0.164 | 0.109 | 0.050 | 0.030 | 0.299 | 8.9E-04 |
| | Attractor Metagene - IFIT3 | Basophils | -0.040 | 0.053 | 0.451 | 0.719 | -0.053 | 0.101 | 0.600 | 0.854 | 0.910 |
| | | Eosinophils | -0.034 | 0.056 | 0.546 | 0.991 | -0.131 | 0.091 | 0.150 | 0.683 | 0.362 |
| | | Lymphocytes | 0.073 | 0.056 | 0.189 | 0.354 | -0.162 | 0.095 | 0.090 | 0.228 | 0.033 |
| | | Neutrophils | 0.031 | 0.055 | 0.572 | 0.794 | 0.048 | 0.107 | 0.658 | 0.964 | 0.892 |
| | | Platelets | -0.075 | 0.049 | 0.131 | 0.655 | -0.110 | 0.087 | 0.209 | 0.729 | 0.723 |
| | | PLR | -0.103 | 0.052 | 0.048 | 0.164 | 0.133 | 0.094 | 0.158 | 0.363 | 0.028 |
| | GP11 Immune IFN | Basophils | -0.005 | 0.016 | 0.760 | 0.828 | -0.012 | 0.024 | 0.606 | 0.854 | 0.796 |
| | | Eosinophils | 0.003 | 0.017 | 0.854 | 0.991 | -0.031 | 0.022 | 0.155 | 0.683 | 0.217 |
| | | Lymphocytes | 0.029 | 0.017 | 0.087 | 0.354 | -0.053 | 0.022 | 0.018 | 0.169 | 3.3E-03 |
| | | Neutrophils | 0.017 | 0.017 | 0.308 | 0.507 | 0.0004 | 0.026 | 0.986 | 0.986 | 0.585 |
| | | Platelets | -0.017 | 0.015 | 0.270 | 0.655 | -0.021 | 0.021 | 0.309 | 0.729 | 0.856 |
| | | PLR | -0.029 | 0.016 | 0.069 | 0.164 | 0.035 | 0.022 | 0.114 | 0.363 | 0.019 |
| | IFN signature 21978456 | Basophils | -0.034 | 0.038 | 0.368 | 0.719 | -0.031 | 0.072 | 0.664 | 0.854 | 0.970 |
| | | Eosinophils | -0.026 | 0.040 | 0.510 | 0.991 | -0.083 | 0.064 | 0.199 | 0.683 | 0.455 |
| | | Lymphocytes | 0.055 | 0.040 | 0.167 | 0.354 | -0.123 | 0.067 | 0.068 | 0.209 | 0.022 |
| | | Neutrophils | 0.016 | 0.040 | 0.693 | 0.794 | 0.036 | 0.076 | 0.642 | 0.964 | 0.817 |
| | | Platelets | -0.053 | 0.035 | 0.136 | 0.655 | -0.080 | 0.062 | 0.202 | 0.729 | 0.708 |
| | | PLR | -0.070 | 0.037 | 0.061 | 0.164 | 0.093 | 0.067 | 0.167 | 0.363 | 0.034 |
| | IFN Cluster 21214954 | Basophils | -0.014 | 0.029 | 0.641 | 0.828 | -0.019 | 0.051 | 0.706 | 0.854 | 0.921 |
| | | Eosinophils | -0.015 | 0.031 | 0.620 | 0.991 | -0.056 | 0.046 | 0.226 | 0.683 | 0.461 |
| | | Lymphocytes | 0.048 | 0.031 | 0.114 | 0.354 | -0.094 | 0.048 | 0.053 | 0.209 | 0.013 |
| | | Neutrophils | 0.024 | 0.030 | 0.425 | 0.626 | 0.021 | 0.055 | 0.705 | 0.964 | 0.957 |

| | | | | | | | | | | | |
|---|---|---|---|---|---|---|---|---|---|---|---|
| | | Platelets | -0.037 | 0.027 | 0.170 | 0.655 | -0.044 | 0.045 | 0.331 | 0.729 | 0.903 |
| | | PLR | -0.062 | 0.028 | 0.030 | 0.164 | 0.069 | 0.048 | 0.151 | 0.363 | 0.019 |
| | IFN signature 19272155 | Basophils | -0.036 | 0.037 | 0.327 | 0.719 | -0.034 | 0.069 | 0.629 | 0.854 | 0.974 |
| | | Eosinophils | -0.030 | 0.039 | 0.447 | 0.991 | -0.081 | 0.062 | 0.195 | 0.683 | 0.483 |
| | | Lymphocytes | 0.052 | 0.039 | 0.179 | 0.354 | -0.121 | 0.065 | 0.065 | 0.209 | 0.022 |
| | | Neutrophils | 0.013 | 0.038 | 0.737 | 0.794 | 0.031 | 0.074 | 0.677 | 0.964 | 0.829 |
| | | Platelets | -0.053 | 0.034 | 0.123 | 0.655 | -0.074 | 0.060 | 0.220 | 0.729 | 0.759 |
| | | PLR | -0.070 | 0.036 | 0.055 | 0.164 | 0.093 | 0.065 | 0.153 | 0.363 | 0.028 |
| | Module3 IFN score | Basophils | -0.021 | 0.035 | 0.540 | 0.796 | -0.034 | 0.063 | 0.597 | 0.854 | 0.864 |
| | | Eosinophils | -0.017 | 0.037 | 0.639 | 0.991 | -0.084 | 0.057 | 0.142 | 0.683 | 0.324 |
| | | Lymphocytes | 0.055 | 0.036 | 0.134 | 0.354 | -0.114 | 0.060 | 0.058 | 0.209 | 0.016 |
| | | Neutrophils | 0.030 | 0.036 | 0.408 | 0.626 | 0.020 | 0.068 | 0.764 | 0.964 | 0.902 |
| | | Platelets | -0.046 | 0.032 | 0.155 | 0.655 | -0.053 | 0.055 | 0.338 | 0.729 | 0.910 |
| | | PLR | -0.068 | 0.034 | 0.045 | 0.164 | 0.098 | 0.059 | 0.098 | 0.363 | 0.014 |
| Leukocyte Subset ES | NK CD56dim cells (ES) | Basophils | 0.005 | 0.014 | 0.715 | 0.828 | -0.003 | 0.032 | 0.923 | 0.943 | 0.813 |
| | | Eosinophils | 0.006 | 0.015 | 0.688 | 0.991 | -0.012 | 0.029 | 0.664 | 0.902 | 0.568 |
| | | Lymphocytes | 0.026 | 0.015 | 0.078 | 0.354 | -0.056 | 0.029 | 0.056 | 0.209 | 0.012 |
| | | Neutrophils | 0.036 | 0.015 | 0.015 | 0.090 | -0.034 | 0.033 | 0.312 | 0.964 | 0.056 |
| | | Platelets | -0.017 | 0.013 | 0.194 | 0.655 | -0.023 | 0.027 | 0.403 | 0.753 | 0.852 |
| | | PLR | -0.012 | 0.014 | 0.369 | 0.450 | 0.042 | 0.029 | 0.145 | 0.363 | 0.088 |
| | Th17 cells (ES) | Basophils | 0.016 | 0.019 | 0.387 | 0.719 | -0.018 | 0.035 | 0.607 | 0.854 | 0.391 |
| | | Eosinophils | 0.005 | 0.020 | 0.784 | 0.991 | 0.004 | 0.032 | 0.901 | 0.939 | 0.970 |
| | | Lymphocytes | 0.017 | 0.020 | 0.398 | 0.530 | -0.006 | 0.034 | 0.870 | 0.900 | 0.570 |
| | | Neutrophils | 0.050 | 0.019 | 0.010 | 0.090 | -0.037 | 0.038 | 0.330 | 0.964 | 0.041 |
| | | Platelets | -0.017 | 0.017 | 0.327 | 0.655 | -0.021 | 0.031 | 0.498 | 0.872 | 0.912 |
| | | PLR | -0.036 | 0.018 | 0.047 | 0.164 | 0.011 | 0.033 | 0.749 | 0.843 | 0.215 |
| | Th2 cells (ES) | Basophils | -0.006 | 0.017 | 0.729 | 0.828 | -0.022 | 0.030 | 0.464 | 0.854 | 0.637 |
| | | Eosinophils | 0.011 | 0.017 | 0.536 | 0.991 | -0.008 | 0.027 | 0.774 | 0.939 | 0.563 |
| | | Lymphocytes | -0.008 | 0.018 | 0.663 | 0.714 | -0.004 | 0.028 | 0.900 | 0.900 | 0.901 |

| | | | | | | | | | | | |
|---|---|---|---|---|---|---|---|---|---|---|---|
| | | Neutrophils | -0.004 | 0.017 | 0.807 | 0.837 | -0.060 | 0.031 | 0.056 | 0.964 | 0.117 |
| | | Platelets | 0.005 | 0.015 | 0.768 | 0.896 | -0.029 | 0.026 | 0.269 | 0.729 | 0.270 |
| | | PLR | 0.026 | 0.016 | 0.106 | 0.223 | 0.007 | 0.028 | 0.803 | 0.843 | 0.544 |
| Macrophage/ Monocyte | Attractor Metagene - G SIGLEC9 | Basophils | 0.089 | 0.071 | 0.215 | 0.719 | 0.029 | 0.104 | 0.782 | 0.854 | 0.634 |
| | | Eosinophils | -0.0004 | 0.075 | 0.995 | 0.995 | -0.015 | 0.093 | 0.876 | 0.939 | 0.906 |
| | | Lymphocytes | 0.116 | 0.075 | 0.125 | 0.354 | -0.085 | 0.097 | 0.380 | 0.532 | 0.101 |
| | | Neutrophils | 0.147 | 0.074 | 0.048 | 0.151 | -0.064 | 0.109 | 0.559 | 0.964 | 0.110 |
| | | Platelets | -0.043 | 0.067 | 0.516 | 0.732 | 0.043 | 0.089 | 0.633 | 0.886 | 0.440 |
| | | PLR | -0.160 | 0.070 | 0.023 | 0.164 | 0.140 | 0.095 | 0.143 | 0.363 | 0.011 |
| | MHC2 signature 21978456 | Basophils | 0.056 | 0.064 | 0.384 | 0.719 | -0.005 | 0.075 | 0.943 | 0.943 | 0.536 |
| | | Eosinophils | 0.078 | 0.067 | 0.243 | 0.991 | 0.055 | 0.068 | 0.413 | 0.723 | 0.809 |
| | | Lymphocytes | 0.104 | 0.067 | 0.122 | 0.354 | -0.073 | 0.070 | 0.303 | 0.532 | 0.069 |
| | | Neutrophils | 0.127 | 0.066 | 0.056 | 0.158 | -0.079 | 0.079 | 0.319 | 0.964 | 0.046 |
| | | Platelets | -0.002 | 0.060 | 0.967 | 0.967 | 0.012 | 0.065 | 0.850 | 0.988 | 0.867 |
| | | PLR | -0.066 | 0.063 | 0.293 | 0.391 | 0.153 | 0.069 | 0.028 | 0.299 | 0.019 |
| T-cell/ Cytotoxic | CD8 T cells (ES) | Basophils | 0.007 | 0.006 | 0.237 | 0.719 | -0.028 | 0.010 | 0.006 | 0.109 | 2.6E-03 |
| | | Eosinophils | -0.004 | 0.006 | 0.535 | 0.991 | -0.010 | 0.009 | 0.266 | 0.683 | 0.561 |
| | | Lymphocytes | 0.007 | 0.006 | 0.267 | 0.440 | -0.031 | 0.009 | 0.001 | 0.037 | 8.3E-04 |
| | | Neutrophils | 0.007 | 0.006 | 0.223 | 0.428 | -0.016 | 0.011 | 0.152 | 0.964 | 0.064 |
| | | Platelets | -0.005 | 0.005 | 0.374 | 0.655 | -0.002 | 0.009 | 0.837 | 0.988 | 0.776 |
| | | PLR | -0.010 | 0.006 | 0.070 | 0.164 | 0.013 | 0.010 | 0.160 | 0.363 | 0.032 |
| | Cytotoxic cells (ES) | Basophils | 0.007 | 0.015 | 0.641 | 0.828 | -0.006 | 0.023 | 0.793 | 0.854 | 0.637 |
| | | Eosinophils | 0.001 | 0.016 | 0.972 | 0.995 | -0.008 | 0.021 | 0.700 | 0.902 | 0.741 |
| | | Lymphocytes | 0.022 | 0.016 | 0.156 | 0.354 | -0.053 | 0.021 | 0.014 | 0.169 | 4.4E-03 |
| | | Neutrophils | 0.031 | 0.015 | 0.043 | 0.151 | 0.004 | 0.025 | 0.856 | 0.970 | 0.357 |
| | | Platelets | -0.013 | 0.014 | 0.357 | 0.655 | -0.006 | 0.020 | 0.760 | 0.967 | 0.787 |
| | | PLR | -0.019 | 0.015 | 0.186 | 0.307 | 0.030 | 0.021 | 0.168 | 0.363 | 0.059 |
| | Eosinophils (ES) | Basophils | 0.002 | 0.007 | 0.722 | 0.828 | 0.008 | 0.014 | 0.580 | 0.854 | 0.727 |
| | | Eosinophils | -0.004 | 0.007 | 0.528 | 0.991 | -0.009 | 0.013 | 0.489 | 0.769 | 0.762 |

| | | | | | | | | | | | |
|---|---|---|---|---|---|---|---|---|---|---|---|
| T-cell/ Cytotoxic | | Lymphocytes | 0.002 | 0.007 | 0.802 | 0.832 | 0.012 | 0.013 | 0.371 | 0.532 | 0.497 |
| | | Neutrophils | 0.003 | 0.007 | 0.676 | 0.794 | 0.010 | 0.015 | 0.512 | 0.964 | 0.673 |
| | | Platelets | -0.007 | 0.006 | 0.229 | 0.655 | 0.000 | 0.012 | 0.973 | 0.988 | 0.569 |
| | | PLR | -0.003 | 0.007 | 0.670 | 0.734 | 0.004 | 0.013 | 0.749 | 0.843 | 0.634 |
| | Macrophages (ES) | Basophils | 0.019 | 0.021 | 0.379 | 0.719 | 0.013 | 0.040 | 0.739 | 0.854 | 0.908 |
| | | Eosinophils | -0.005 | 0.022 | 0.807 | 0.991 | -0.013 | 0.036 | 0.709 | 0.902 | 0.850 |
| | | Lymphocytes | 0.039 | 0.022 | 0.080 | 0.354 | -0.030 | 0.037 | 0.419 | 0.533 | 0.111 |
| | | Neutrophils | 0.051 | 0.022 | 0.019 | 0.090 | -0.023 | 0.042 | 0.590 | 0.964 | 0.118 |
| | | Platelets | -0.004 | 0.020 | 0.846 | 0.911 | -0.018 | 0.034 | 0.591 | 0.883 | 0.712 |
| | | PLR | -0.038 | 0.021 | 0.067 | 0.164 | 0.043 | 0.037 | 0.239 | 0.459 | 0.053 |
| | Neutrophils (ES) | Basophils | 0.021 | 0.016 | 0.192 | 0.719 | 0.021 | 0.032 | 0.501 | 0.854 | 0.981 |
| | | Eosinophils | 0.020 | 0.017 | 0.233 | 0.991 | -0.002 | 0.029 | 0.939 | 0.939 | 0.505 |
| | | Lymphocytes | 0.038 | 0.016 | 0.023 | 0.328 | -0.023 | 0.030 | 0.446 | 0.543 | 0.076 |
| | | Neutrophils | 0.047 | 0.016 | 0.004 | 0.090 | -0.005 | 0.033 | 0.875 | 0.970 | 0.156 |
| | | Platelets | -0.002 | 0.015 | 0.905 | 0.938 | 0.014 | 0.027 | 0.598 | 0.883 | 0.602 |
| | | PLR | -0.035 | 0.015 | 0.024 | 0.164 | 0.019 | 0.029 | 0.521 | 0.729 | 0.104 |
| | NK cells (ES) | Basophils | 0.004 | 0.006 | 0.462 | 0.719 | -0.008 | 0.012 | 0.484 | 0.854 | 0.339 |
| | | Eosinophils | -0.001 | 0.006 | 0.840 | 0.991 | -0.011 | 0.010 | 0.312 | 0.683 | 0.440 |
| | | Lymphocytes | 0.0005 | 0.006 | 0.937 | 0.937 | -0.017 | 0.011 | 0.109 | 0.235 | 0.149 |
| | | Neutrophils | 0.002 | 0.006 | 0.713 | 0.794 | -0.003 | 0.012 | 0.792 | 0.964 | 0.690 |
| | | Platelets | -0.006 | 0.005 | 0.282 | 0.655 | -0.009 | 0.010 | 0.394 | 0.753 | 0.808 |
| | | PLR | -0.003 | 0.006 | 0.566 | 0.660 | 0.009 | 0.011 | 0.415 | 0.646 | 0.322 |
| | T helper cells (ES) | Basophils | 0.008 | 0.008 | 0.347 | 0.719 | -0.040 | 0.015 | 0.008 | 0.109 | 5.0E-03 |
| | | Eosinophils | -0.004 | 0.009 | 0.630 | 0.991 | -0.025 | 0.013 | 0.060 | 0.683 | 0.183 |
| | | Lymphocytes | 0.007 | 0.009 | 0.428 | 0.545 | -0.012 | 0.014 | 0.413 | 0.533 | 0.266 |
| | | Neutrophils | 0.010 | 0.008 | 0.253 | 0.442 | -0.024 | 0.016 | 0.136 | 0.964 | 0.063 |
| | | Platelets | -0.002 | 0.008 | 0.761 | 0.896 | -0.013 | 0.013 | 0.306 | 0.729 | 0.463 |
| | | PLR | -0.003 | 0.008 | 0.682 | 0.734 | 0.014 | 0.014 | 0.309 | 0.541 | 0.276 |
| | | Basophils | 0.011 | 0.011 | 0.318 | 0.719 | -0.045 | 0.023 | 0.056 | 0.519 | 0.030 |

| | | | | | | | | | | | |
|---|---|---|---|---|---|---|---|---|---|---|---|
| T-cell/ Cytotoxic | T-CM cells (ES) | Eosinophils | 0.014 | 0.011 | 0.218 | 0.991 | -0.031 | 0.021 | 0.141 | 0.683 | 0.059 |
| | | Lymphocytes | 0.014 | 0.011 | 0.218 | 0.381 | -0.027 | 0.022 | 0.236 | 0.471 | 0.106 |
| | | Neutrophils | 0.015 | 0.011 | 0.191 | 0.428 | 0.002 | 0.025 | 0.948 | 0.983 | 0.640 |
| | | Platelets | 0.007 | 0.010 | 0.462 | 0.722 | -0.026 | 0.021 | 0.218 | 0.729 | 0.152 |
| | | PLR | -0.002 | 0.011 | 0.869 | 0.869 | 0.012 | 0.022 | 0.588 | 0.748 | 0.575 |
| | T-EM cells (ES) | Basophils | 0.008 | 0.007 | 0.250 | 0.719 | -0.008 | 0.019 | 0.657 | 0.854 | 0.418 |
| | | Eosinophils | 0.001 | 0.007 | 0.927 | 0.995 | -0.028 | 0.017 | 0.101 | 0.683 | 0.120 |
| | | Lymphocytes | 0.019 | 0.007 | 0.006 | 0.178 | -0.017 | 0.018 | 0.331 | 0.532 | 0.055 |
| | | Neutrophils | 0.018 | 0.007 | 0.009 | 0.090 | 0.011 | 0.020 | 0.584 | 0.964 | 0.735 |
| | | Platelets | -0.005 | 0.006 | 0.464 | 0.722 | -0.026 | 0.016 | 0.110 | 0.729 | 0.217 |
| | | PLR | -0.008 | 0.007 | 0.241 | 0.353 | -0.003 | 0.017 | 0.843 | 0.843 | 0.819 |
| | Tfh cells (ES) | Basophils | -0.007 | 0.008 | 0.402 | 0.719 | -0.020 | 0.019 | 0.314 | 0.854 | 0.547 |
| | | Eosinophils | -0.007 | 0.009 | 0.404 | 0.991 | 0.017 | 0.018 | 0.341 | 0.683 | 0.220 |
| | | Lymphocytes | -0.005 | 0.009 | 0.538 | 0.602 | -0.016 | 0.018 | 0.380 | 0.532 | 0.594 |
| | | Neutrophils | 0.003 | 0.009 | 0.693 | 0.794 | 0.007 | 0.021 | 0.723 | 0.964 | 0.859 |
| | | Platelets | 0.005 | 0.008 | 0.549 | 0.732 | -0.018 | 0.017 | 0.297 | 0.729 | 0.230 |
| | | PLR | 0.002 | 0.008 | 0.790 | 0.819 | 0.004 | 0.018 | 0.813 | 0.843 | 0.914 |
| | Th1 cells (ES) | Basophils | -0.003 | 0.010 | 0.769 | 0.828 | -0.008 | 0.017 | 0.645 | 0.854 | 0.797 |
| | | Eosinophils | -0.008 | 0.010 | 0.455 | 0.991 | -0.013 | 0.015 | 0.387 | 0.723 | 0.758 |
| | | Lymphocytes | 0.006 | 0.010 | 0.522 | 0.602 | -0.025 | 0.015 | 0.104 | 0.235 | 0.086 |
| | | Neutrophils | 0.004 | 0.010 | 0.723 | 0.794 | -0.014 | 0.017 | 0.412 | 0.964 | 0.373 |
| | | Platelets | -0.008 | 0.009 | 0.353 | 0.655 | -0.001 | 0.014 | 0.970 | 0.988 | 0.646 |
| | | PLR | -0.011 | 0.009 | 0.252 | 0.353 | 0.003 | 0.015 | 0.837 | 0.843 | 0.439 |
| TGF- β | TGFB PCA 17349583 | Basophils | 0.031 | 0.033 | 0.352 | 0.719 | 0.029 | 0.073 | 0.686 | 0.854 | 0.982 |
| | | Eosinophils | 0.007 | 0.035 | 0.833 | 0.991 | -0.035 | 0.066 | 0.596 | 0.879 | 0.571 |
| | | Lymphocytes | 0.051 | 0.035 | 0.151 | 0.354 | -0.021 | 0.068 | 0.755 | 0.846 | 0.348 |
| | | Neutrophils | 0.076 | 0.035 | 0.028 | 0.113 | -0.086 | 0.076 | 0.261 | 0.964 | 0.053 |
| | | Platelets | -0.008 | 0.031 | 0.802 | 0.898 | -0.001 | 0.063 | 0.988 | 0.988 | 0.922 |
| | | PLR | -0.040 | 0.033 | 0.227 | 0.353 | 0.078 | 0.067 | 0.246 | 0.459 | 0.114 |

**Supplementary Figure S1: Overview of glioma study populations.** For each study analyses were restricted to participants of predominantly European ancestry. Genome-wide association analyses from each study were combined using fixed effects meta-analysis and the results summary statistics were used for all Mendelian randomization analyses. For each molecular subtype, the number of mortality events is reported in brackets. Chromosome 1p/19q co-deletion typically does not occur in IDH wildtype tumors, therefore information on this molecular marker is not always present or assessed for this suptype.

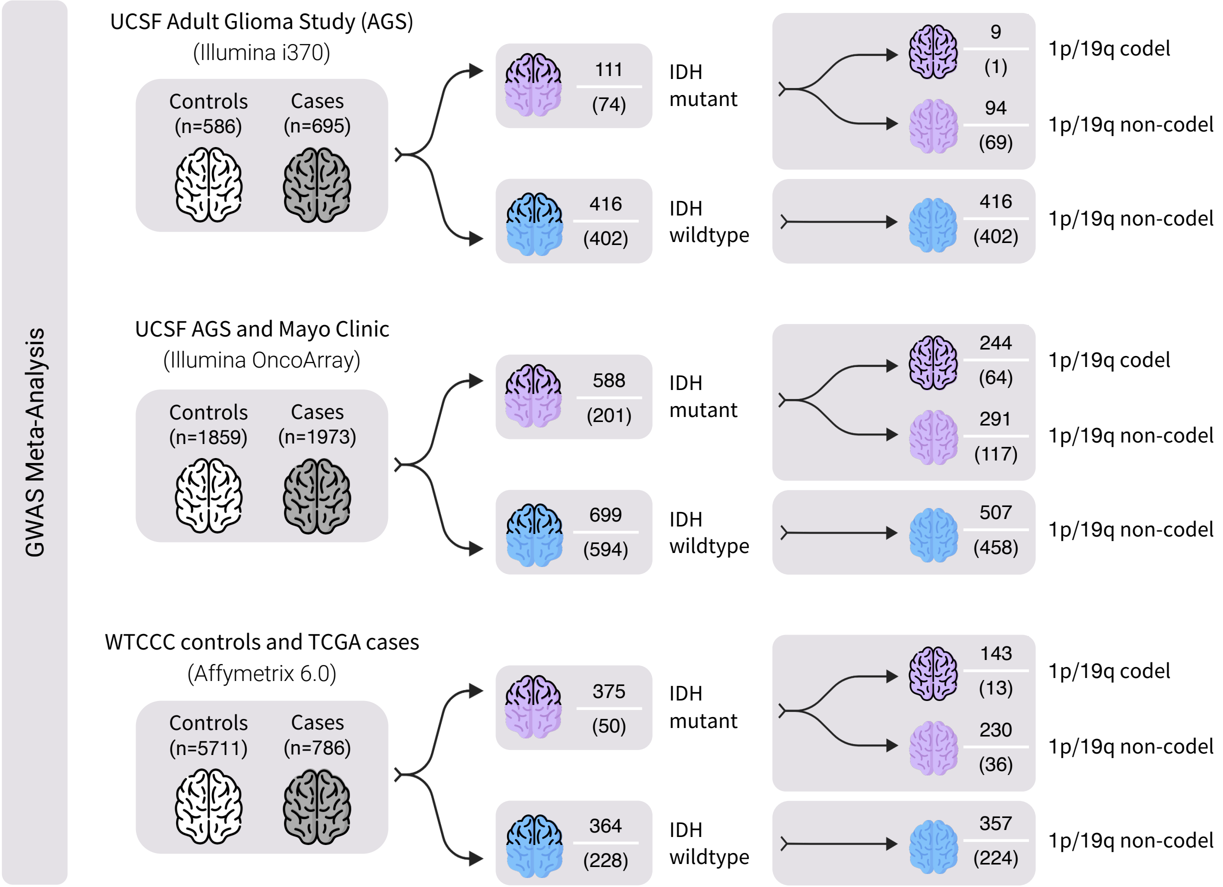

**Supplementary Figure S2: Effects on gene expression in brain tissues.** Prevalence of expression quantitative trait loci (eQTL) in GTEx v8 brain tissues among genetic instruments for blood cell traits and eGenes in regions linked to glioma susceptibility.

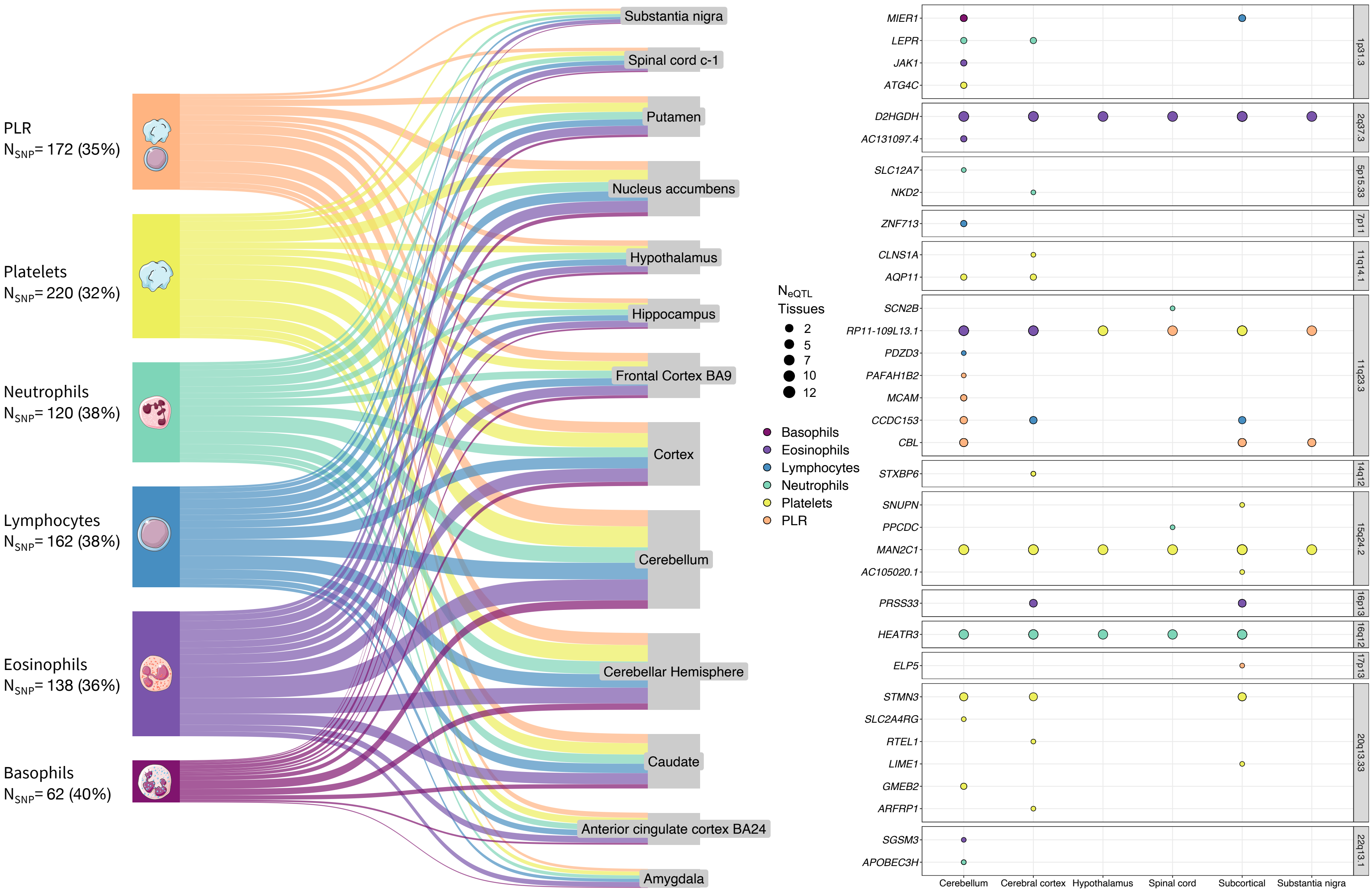

**Supplementary Figure S3: Regional association and z-score plots for colocalized signals.** In each plot the LD is calculated with respect to the shared fine-mapped variant, indicated by the black diamond. For chromosome 20q13.33, the lead telomere length variant (rs13038527) is also shown, although the signal for telomere length did not colocalize.

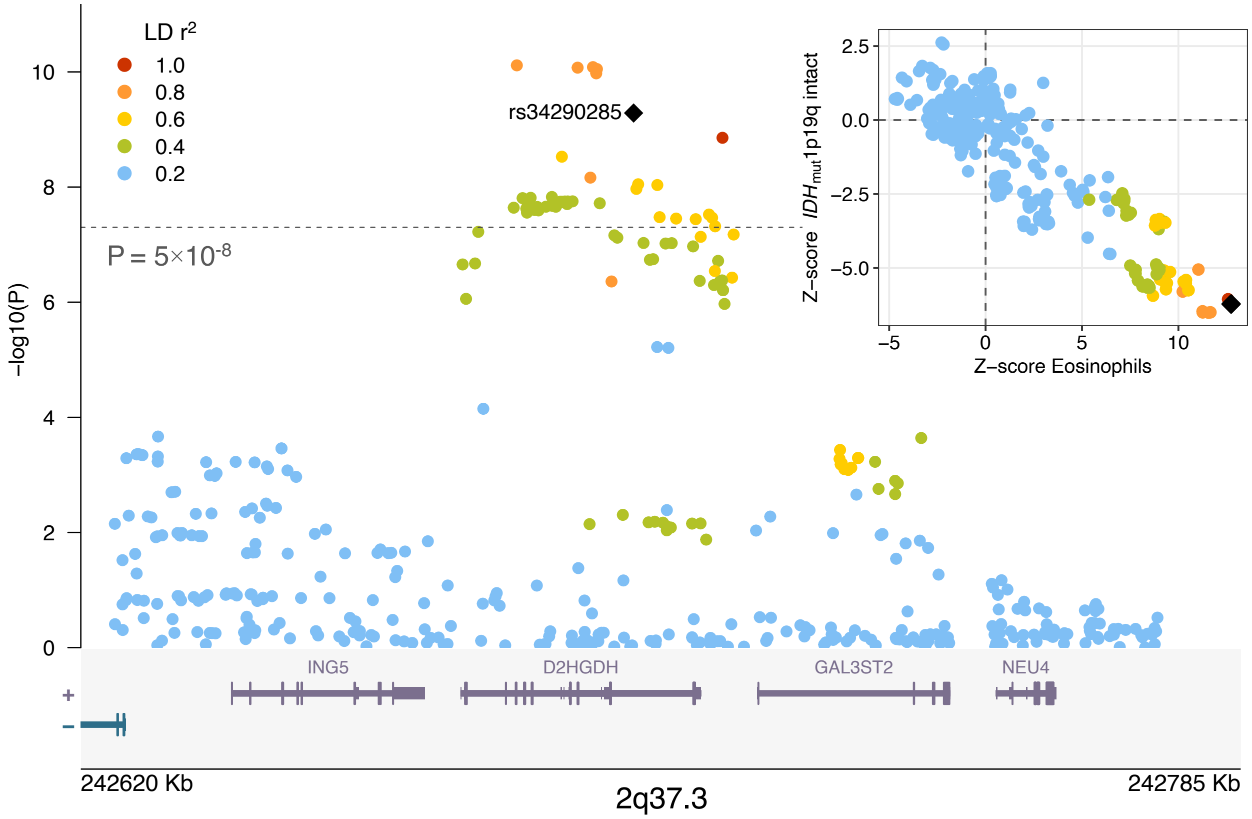


2q37.3

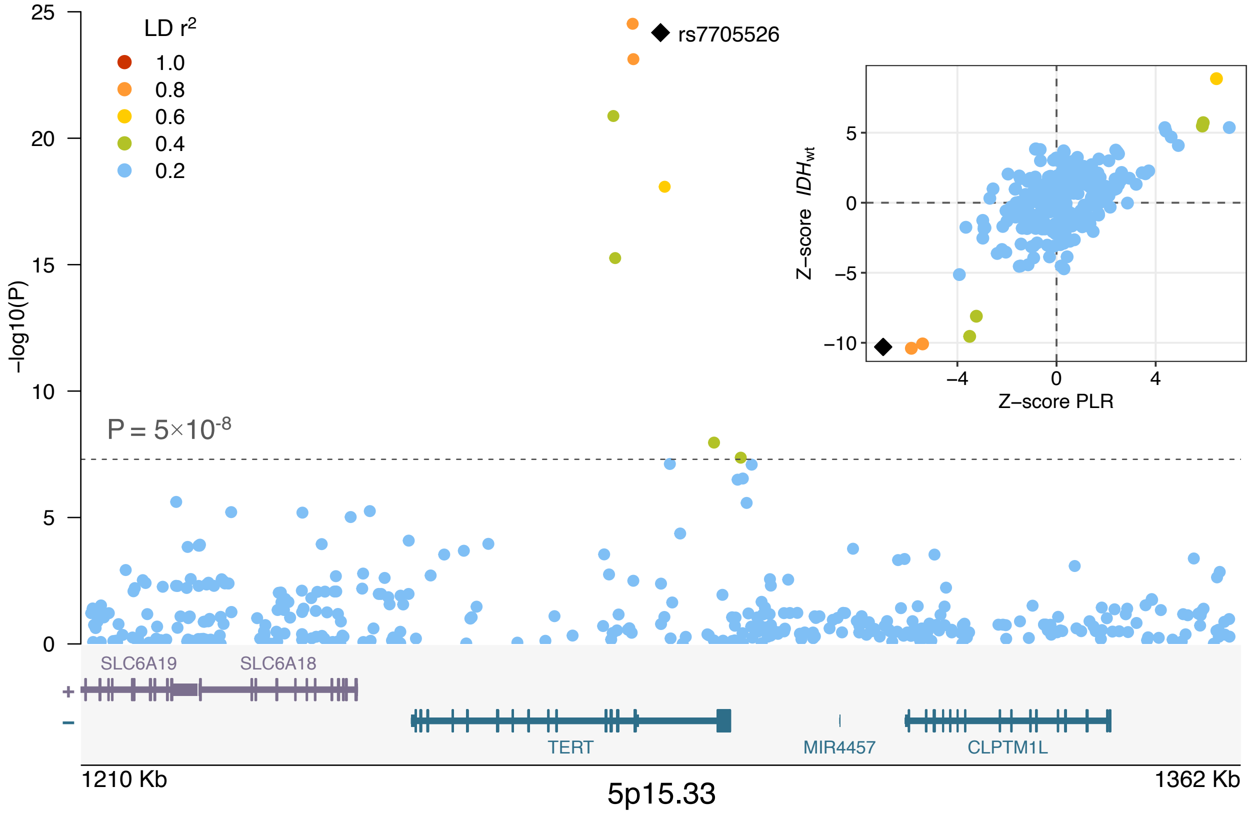


5p15.33

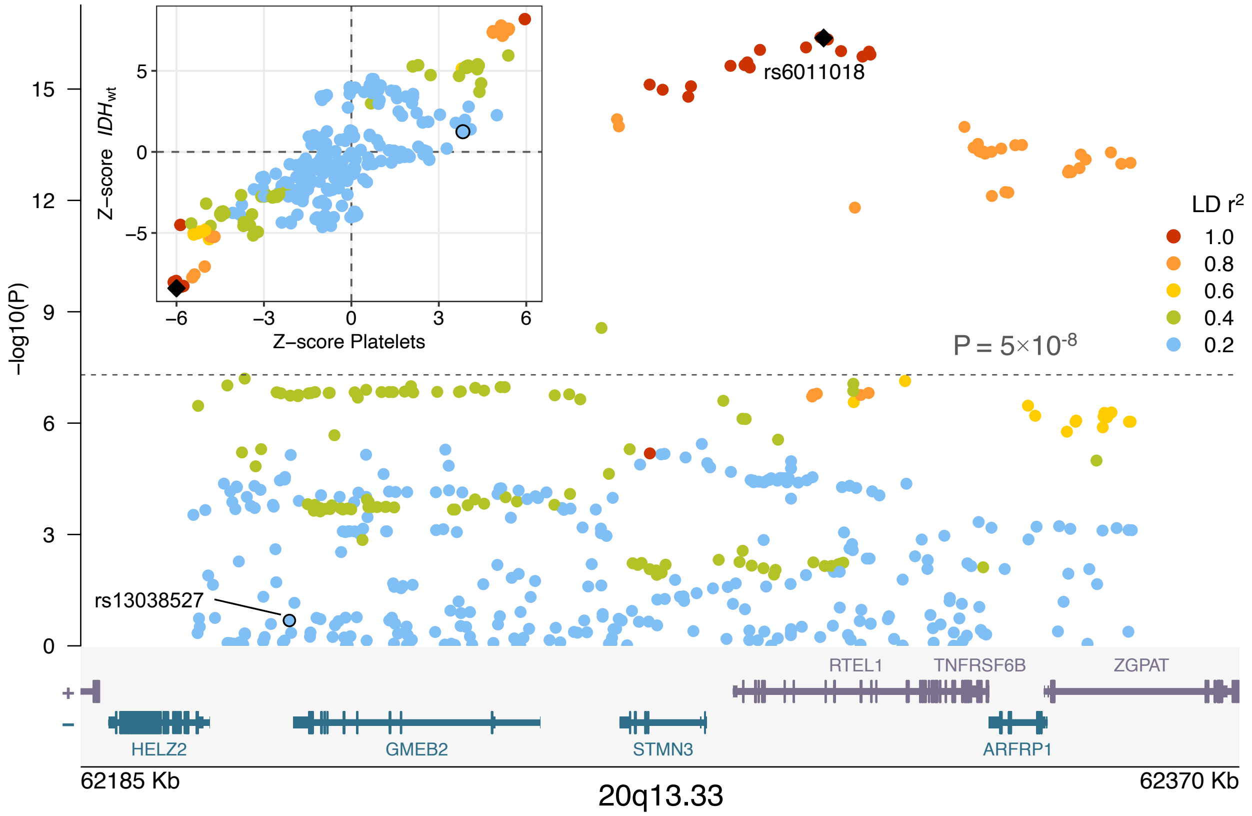

-log10(P)
Z-score IDHwt
Z–score Platelets
rs6011018
rs13038527
P = 5×10-8
LD r2
1.0
0.8
0.6
0.4
0.2
RTEL1
TNFRSF6B
ZGPAT
HELZ2
GMEB2
STMN3
ARFRP1
62185 Kb
20q13.33
62370 Kb
